# Resting-state EEG activity as a Biomarker and Treatment Target in Depression: A Systematic Review and Meta-analysis

**DOI:** 10.1101/2025.10.22.25338525

**Authors:** Henrik Heitmann, Jean-Francois Siani, Paul Theo Zebhauser, Peter Henningsen, Stefan Leucht, Josef Priller, Markus Ploner

## Abstract

Depression is a highly prevalent and disabling disorder affecting approximately 5% of the adult population worldwide. Despite its impact, the underlying pathophysiology remains insufficiently understood, and current treatments are only partially effective. Brain-based biomarkers offer promise for clarifying mechanisms of depression and guiding novel treatment approaches, including neuromodulation. EEG is particularly attractive for this purpose due to its wide availability, cost-effectiveness, and potential for direct neuromodulatory targeting.

We conducted a PROSPERO-registered systematic review in accordance with PRISMA guidelines to assess resting-state EEG biomarkers in adult patients with depression, diagnosed according to DSM-IV/V or ICD-10/11. Included studies reported cross-sectional or correlational data on quantitative EEG measures such as power, cordance, peak frequency, and alpha asymmetry. Semiquantitative analyses using modified albatross plots and meta-analyses were performed. Study quality was assessed with a modified Newcastle-Ottawa Scale.

Fifty-two studies met the inclusion criteria. Findings indicated increased low-frequency (delta, theta) and high-frequency (beta, gamma) power, and left frontal alpha asymmetry in depressed patients compared to healthy controls. Meta-analysis confirmed a significant increase in beta power. However, results regarding disease severity correlations and data on peak alpha frequency and cordance were insufficient for interpretation. Risk of bias across studies was high.

Our results support increased beta and potentially also theta oscillations and alpha asymmetry as candidate diagnostic EEG biomarkers for depression. These oscillations may reflect disrupted corticolimbic control and reward processing and partially overlap with mechanisms implicated in chronic pain and fatigue. Further investigation is warranted into their potential as diagnostic tools and neuromodulatory treatment targets.

## Introduction

Depression is a highly prevalent and disabling disorder affecting around 5% of the adult population worldwide, imposing a significant burden on societies and healthcare systems (Collaborators 2022, Shorey, Ng et al. 2022). Despite considerable research, its pathophysiology is not fully clear, and treatments are often insufficient (Nemeroff 2020).

The diagnosis of depression is established according to disease classifications, with the Diagnostic and Statistical Manual of Mental Disorders (DSM) and the International Statistical Classification of Diseases and Health Related Problems (ICD) being the most established. Diagnosis and monitoring of depression according to these frameworks rely on questionnaires and interviews. These subjective assessments might be complemented and extended by objective biomarkers for the diagnosis and monitoring of depression. According to the National Institutes of Health (NIH) Biomarkers, EndpointS, and other Tools (BEST) classification, biomarkers can fulfill different functions. Diagnostic biomarkers might allow the differentiation of patients with depression from healthy participants, and monitoring biomarkers may enable evaluation and tracking of depression severity, including treatment responses, which might be particularly valuable (FDA-NIH Biomarker Working Group 2016). Since depression is associated with aberrant brain activity (Muller, Cieslik et al. 2017), biomarkers based on brain activity might help to understand the pathophysiology and to develop targeted treatment approaches, e.g., using neuromodulation techniques (de Aguiar Neto and Rosa 2019, Newson and Thiagarajan 2019, Marwaha, Palmer et al. 2023). Using resting-state electroencephalography (EEG) to detect such biomarkers is appealing since it is broadly available, cost-effective, and potentially scalable (de Aguiar Neto and Rosa 2019). Correspondingly, an increasing number of studies have investigated the potential of various EEG parameters, e.g., as diagnostic biomarkers and predictors of treatment response in depression (Olbrich, van Dinteren et al. 2015). Moreover, recent neuromodulatory treatment approaches, such as non-invasive brain stimulation, allow for direct targeting of brain activity captured by EEG, thereby further underlining the translational potential of EEG findings (de Aguiar Neto and Rosa 2019). Previous systematic reviews point towards altered EEG power, asymmetry, and connectivity in patients with depression, with an increase in low-frequency band power and alpha asymmetry with relatively increased left-sided activity in patients compared to healthy participants being most consistently described (van der Vinne, Vollebregt et al. 2017, de Aguiar Neto and Rosa 2019, Newson and Thiagarajan 2019, Miljevic, Bailey et al. 2023, Luo, Tang et al. 2025). While these studies provide valuable insights, the wide variety of EEG parameters investigated often hindered quantitative analysis and interpretation of results (e.g., meta-analysis) (Tsai, Li et al. 2023). Furthermore, the heterogeneity of study populations included, often lacking clear diagnostic criteria for depression, makes it challenging to derive clinically meaningful conclusions and approaches.

To address these challenges, the present study aims to provide a comprehensive overview of potential diagnostic and monitoring EEG biomarkers that could serve as potential treatment targets for depression. To this end, the relationship between depression, as defined by DSM-IV/DSM-5 and ICD-10/ICD-11, and well-established quantitative oscillation-related EEG parameters, including band-specific power, alpha asymmetry, peak alpha frequency, and cordance, is investigated. Alpha asymmetry refers to an imbalance between left and right hemisphere frontal alpha band power, which has been observed in depression. Mechanistically, it has been hypothesized to reflect conflicting appetitive and aversive behaviour (van der Vinne, Vollebregt et al. 2017). The peak alpha frequency (PAF) denotes the peak frequency of the power spectrum in the alpha frequency band and has been related to the individual responses to pharmacological and brain stimulation treatments in depression (Voetterl, Sack et al. 2023). Cordance is a measure of regional brain activity combining absolute and relative EEG power (Leuchter 1994, de la Salle, Jaworska et al. 2020). It has been suggested to be particularly closely related to brain metabolism and implicated in treatment response prediction to antidepressant medications and interventions, including brain stimulation (de la Salle, Jaworska et al. 2020). Focusing on such frequently analyzed EEG parameters enables a standardized and reproducible approach with high translational applicability. This holds the potential to contribute to a better pathophysiological understanding of depression, aiding its diagnosis and treatment.

## Methods

The present study was conducted and is reported following the Preferred Reporting Items for Systematic Reviews and Meta-Analyses (PRISMA) (Page, McKenzie et al. 2021). The study protocol was preregistered on PROSPERO (Identifier CRD 42024492853). The process of deduplication, screening of title and abstract, as well as full-text review and data extraction, were performed using the software Covidence (Covidence 2021). The present follows the methodology of previous systematic reviews performed for chronic pain, fatigue and recently migraine (Heitmann, Zebhauser et al. 2023, Zebhauser, Hohn et al. 2023, Zebhauser, Heitmann et al. 2024).

### Search strategy

The databases MEDLINE, PubMed Central, and Bookshelf (through PubMed), Web of Science Core Collection (through Web of Science), and EMBASE (through Ovid) were searched. The search strings used comprised combinations of depression and EEG, using Boolean operators and truncations, and can be found in detail for each database in the supplementary material. Databases were searched from their inception dates until 22 February and again on 5 September 2024. No language restrictions were applied. Additionally, reference mining of recent reviews on EEG in depression (de Aguiar Neto and Rosa 2019, Newson and Thiagarajan 2019, Miljevic, Bailey et al. 2023) and included studies were performed.

### Study selection

For detailed inclusion and exclusion criteria, please see Table 1. In summary, cross-sectional and correlational data from peer-reviewed studies measuring quantitative EEG activity measures (power, asymmetry, peak frequency, and cordance) in awake resting-state adult human patients suffering from depression according to DSM IV/V or ICD 10/11 were included. Studies involving people with other severe neuropsychiatric disorders were excluded.

**Table 1.** Inclusion and exclusion criteria.

| Inclusion (if all apply) | Exclusion (if any applies) |
| --- | --- |
| Depression (according to DSM IV/V or ICD 10/11) as the primary condition studied | Other severe neuropsychiatric comorbidities incl. substance-induced mood-disorders |
| Wake resting-state EEG recording in adult humans |  |
| At least one of the following quantitative EEG measures reported<br>- Band-specific power<br>- Band-specific asymmetry<br>- Peak-frequency<br>- Cordance |  |
| Cross-sectional comparison with healthy control group or correlation with disease severity |  |
| Published and peer-reviewed studies | Review articles and case studies |

### Record screening, full-text review, and data extraction

Titles and abstracts were independently screened by two authors blinded to each other’s decision. In case of disagreement, conflicts were discussed and resolved. This was performed likewise for full-text review. One author performed the extraction, and another author verified the results. The data extracted comprised general study information, including participant details, EEG recording specifications, and outcome measures.

For cross-sectional group comparisons of EEG features (t-tests and Mann-Whitney-U-tests), the following parameters were extracted for meta-analysis: Means and standard deviations (SDs), t-values, U-values, and p-values.

For correlations of disease severity with EEG features (Pearson or Spearman correlations), r-/rho- and p-values were extracted.

If necessary, algebraic recalculation of means, SDs, and effect sizes was implemented following recent recommendations (Covidence 2021). Data was extracted from figures whenever essential and possible. We contacted study authors to retrieve statistics whenever algebraic recalculation was mathematically impossible. In the case of multiple comparisons reported for one EEG feature (e.g., a study had analyzed several regions of interest for oscillatory theta power), the most significant effect was selected for further analysis. If imprecise p-values for significant findings were reported (for example, “*p*<*0.01*”), we used the closest decimal value for extraction (“*p*=*0.009*”). If p-values for non-significant findings were reported (e.g., “*p*>*0.05*”), we chose not to extract the nearest decimal since a valid approximation of the measured effect could not be guaranteed. For studies looking at alpha asymmetry, correction for the directionality of effects (right-left) was performed in line with previous systematic reviews (Luo, Tang et al. 2025). Thus, negative values point towards increased left-sided oscillatory alpha activity and vice versa.

### Data synthesis

A multi-step approach was used for data synthesis, considering the number and quality of studies.

First, semiquantitative analyses were performed using modified albatross plots and vote counting, as previously reported for all comparisons and correlations of interest (Heitmann, Zebhauser et al. 2023, Zebhauser, Hohn et al. 2023). For correlations, albatross plots show p-values for negative correlations with disease severity on the left side of the panel and positive correlations on the right side, respectively. This allowed for the inclusion of studies that only provided imprecise p-values, as reported above. Second, a meta-analysis was performed if k>4 studies reporting the necessary precise study information were available for the corresponding group comparison or correlation, following recent recommendations on the minimum number of studies needed for random-effect meta-analysis (Jackson and Turner 2017). Meta-analysis was performed using R Version 4.1.2 (R Core Team 2021) with the *metafor* package (Viechtbauer 2010). Random-effect models were chosen due to anticipated significant between-study heterogeneity in (i) EEG data acquisition and analysis and (ii) clinical characteristics of study participants. Heterogeneity was evaluated with Cochran’s *Q* (*p*<0.05 indicating heterogeneity) and *I^2^* (values of 25%, 50%, and 75% representing low, moderate, and high heterogeneity, respectively). Funnel plots and Egger’s tests were used to assess publication bias.

For group comparisons, Hedges’ g was used to compare EEG features between groups due to the small sample sizes of studies. To that end, for studies using parametric statistical tests (t-tests), effect sizes were calculated directly from means and SDs, p-values, and sample sizes. For studies using non-parametric tests (Mann-Whitney-U-tests), eta-squared was calculated as an effect size estimate (Fritz, Morris et al. 2012) and converted to Hedges’ g using the *esc* package in R as previously reported (Zebhauser, Heitmann et al. 2024). For correlation studies, r- and rho-values were used. Narrative data synthesis was used for the remaining studies.

### Risk of bias and quality assessment

The risk of bias (RoB) and study quality was assessed using a modified Newcastle-Ottawa-Scale in terms of “selection of study participants,” “comparability/ confounders,” and “outcome data” (see supplemental material). In the original scale version, stars were awarded for individual domains, whereas the present version used rated items as “high” or “low” RoB for a more straightforward interpretation.

### Missing data and full texts

Corresponding authors were contacted up to two times via email to request missing data or inaccessible full texts. Data/full texts were considered unavailable if no reply was received four weeks after the second contact attempt.

## Results

### Study selection and characteristics

The database searches yielded 2424 records after deduplication. Screening identified 390 studies, of which 52 were finally included (Dharmadhikari, Jaiswal et al., Knott, Mahoney et al. 2000, Knott, Mahoney et al. 2001, Pizzagalli, Nitschke et al. 2002, Allen, Urry et al. 2004, Morgan, Witte et al. 2005, Strelets, Garakh et al. 2007, Korb, Cook et al. 2008, Putnam and McSweeney 2008, Allen and Cohen 2010, Farahbod, Cook et al. 2010, Kemp, Griffiths et al. 2010, Saletu, Anderer et al. 2010, Begić, Popović-Knapić et al. 2011, Segrave, Cooper et al. 2011, Jaworska, Blier et al. 2012, Gold, Fachner et al. 2013, Plante, Goldstein et al. 2013, Cook, Hunter et al. 2014, Escolano, Navarro-Gil et al. 2014, Quinn, Rennie et al. 2014, Arns, Etkin et al. 2015, Arns, Gordon et al. 2015, Cantisani, Koenig et al. 2015, Tas, Cebi et al. 2015, Arns, Bruder et al. 2016, Roh, Park et al. 2016, Scanlon, Jain et al. 2016, Mumtaz, Xia et al. 2017, Arikan, Gunver et al. 2019, Kim, Oh et al. 2019, Koo-Poeggel, Berger et al. 2019, Soukhtanlou, Rostami et al. 2019, Čukić, Stokić et al. 2020, Das and Yadav 2020, Roh, Kim et al. 2020, Hill, Zomorrodi et al. 2021, Lin, Chen et al. 2021, Kesebir, Yosmaoglu et al. 2022, Liu, Liu et al. 2022, Liu, Liu et al. 2022, Wu, Zhong et al. 2022, Huang, Yi et al. 2023, Jang, Kim et al. 2023, Jiang, Huang et al. 2023, Lin, Du et al. 2023, Marcu, Szekely-Copîndean et al. 2023, Xia, Wang et al. 2023, Liu, Zhang et al. 2024, Zeng, Lao et al. 2024). The PRISMA flow diagram of study selection and exclusion reasons at the different levels is shown in Figure 1.

**Figure 1:**
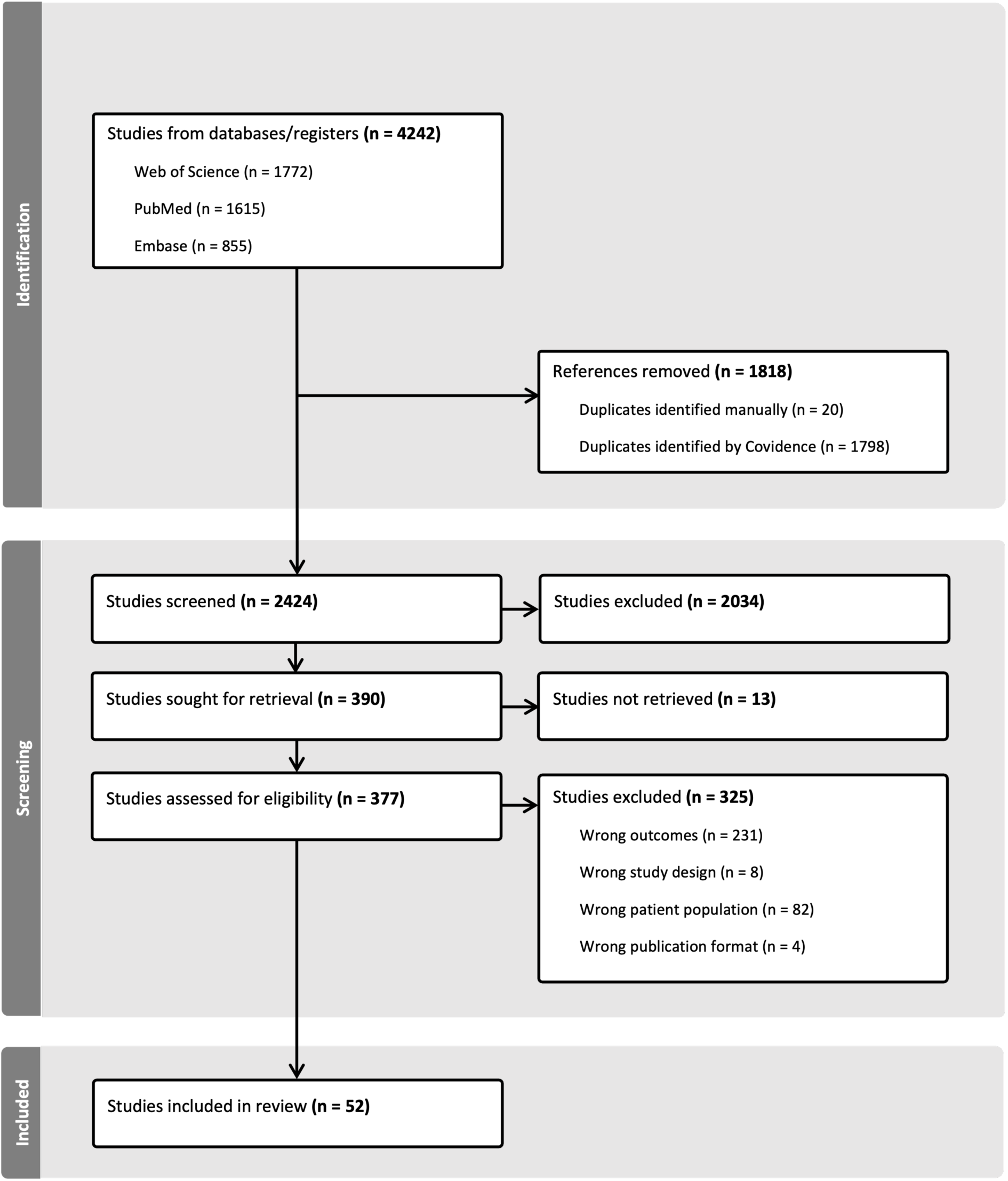
PRISMA-Flowchart.

### Risk of bias and study quality

An overview of the results of the risk of bias (RoB) assessment is provided in Figure 2. The individual studies’ scoring can be found in the supplementary material. In the “selection of participants”-domain, the most significant RoB was related to “case representativeness”, as half of the studies did not describe their sampling strategy in detail. Case definition was unproblematic given the strict inclusion criteria for studies regarding diagnostic criteria of patient samples. In the “comparability/confounders”-domain, there was considerable RoB regarding the lack of controlling for anxiety as the most frequent comorbidity of depression and other potential confounding factors such as other comorbidity or medication intake. For the “outcome data”-domain, a high RoB was obtained related to a lack of blinding for the outcome assessment and a partially limited description of the statistical testing applied.

**Figure 2:**
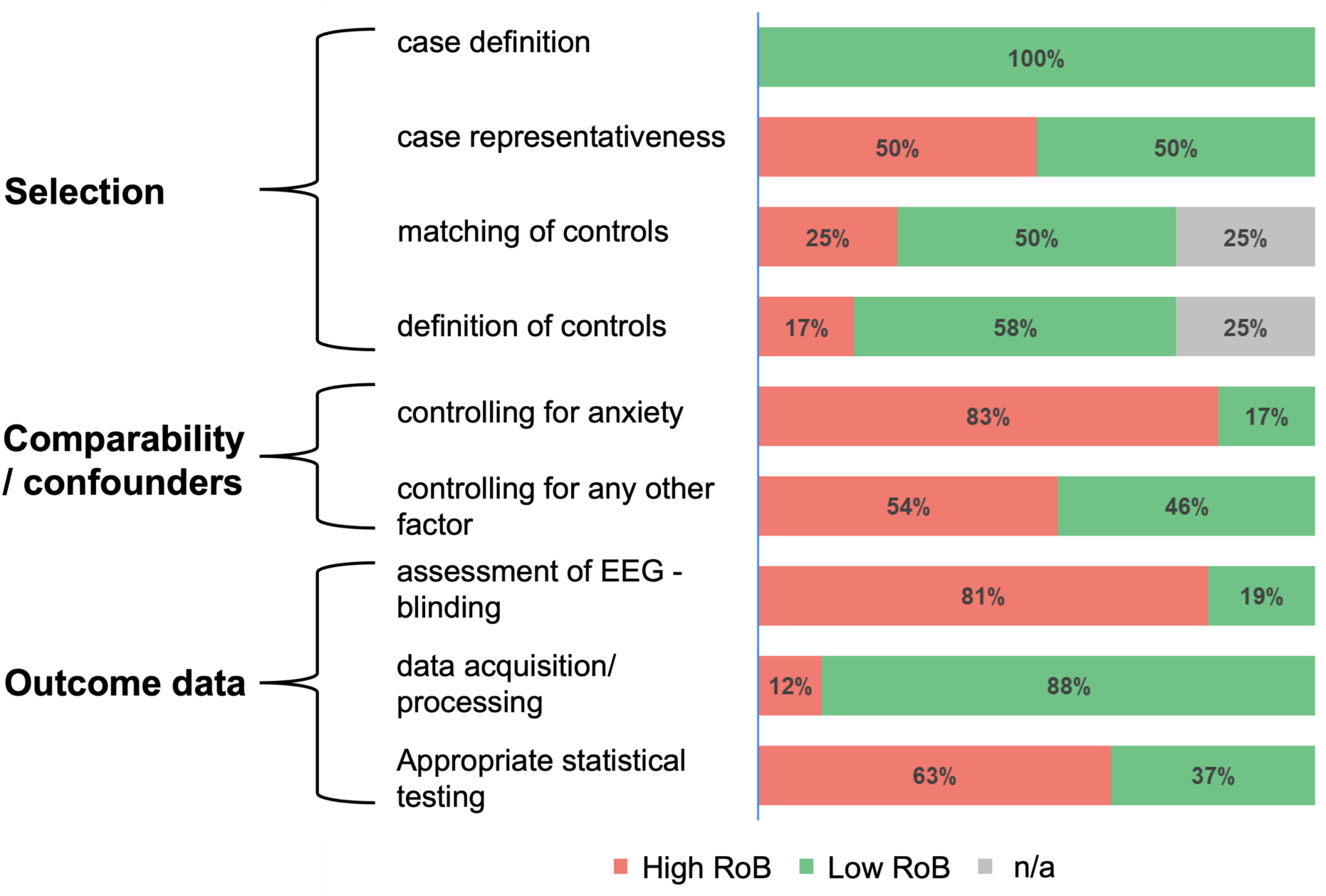
Summary of the Risk of bias (RoB) assessment.

Overall, there was considerable RoB (>50% of studies) in 5/9 domains, with a lack of controlling for relevant comorbidity and non-blinded EEG assessment being the most prominent. For a detailed RoB on the study level, please see Supplementary Figure S1.

### Data Synthesis

Two types of data were included in the analyses. Data from cross-sectional studies comparing EEG parameters in patients with depression compared to healthy participants, as well as data from studies reporting the correlation of depression severity with EEG parameters. Data analysis and synthesis followed a two-step approach in the case of four or more studies providing data for the respective EEG parameters. First, as previously reported, semiquantitative data analyses were performed using modified albatross and vote counting (Heitmann, Zebhauser et al. 2023, Zebhauser, Hohn et al. 2023). Second, a meta-analysis was performed if four or more studies provided sufficient data (Zebhauser, Heitmann et al. 2024). For less frequently reported EEG parameters, narrative synthesis was performed.

#### Group comparisons (depression vs. healthy participants)

A total of 33 studies performed cross-sectional group comparisons of one or more EEG parameters in patients with depression compared to healthy participants. Of those, 26 studies reported a comparison of band-specific EEG power (Dharmadhikari, Jaiswal et al., Knott, Mahoney et al. 2001, Morgan, Witte et al. 2005, Strelets, Garakh et al. 2007, Korb, Cook et al. 2008, Begić, Popović-Knapić et al. 2011, Jaworska, Blier et al. 2012, Zeng, Shen et al. 2012, Plante, Goldstein et al. 2013, Cook, Hunter et al. 2014, Arns, Etkin et al. 2015, Mumtaz, Xia et al. 2017, Arikan, Gunver et al. 2019, Soukhtanlou, Rostami et al. 2019, Čukić, Stokić et al. 2020, Das and Yadav 2020, Hill, Zomorrodi et al. 2021, Lin, Chen et al. 2021, Liu, Liu et al. 2022, Liu, Liu et al. 2022, Wu, Zhong et al. 2022, Huang, Yi et al. 2023, Jang, Kim et al. 2023, Jiang, Huang et al. 2023, Lin, Du et al. 2023, Xia, Wang et al. 2023). Comparison of alpha asymmetry (Luo, Tang et al. 2025) (AA) was reported in 14 studies (Dharmadhikari, Jaiswal et al., Knott, Mahoney et al. 2001, Allen and Cohen 2010, Kemp, Griffiths et al. 2010, Segrave, Cooper et al. 2011, Jaworska, Blier et al. 2012, Cantisani, Koenig et al. 2015, Arns, Bruder et al. 2016, Mumtaz, Xia et al. 2017, Koo-Poeggel, Berger et al. 2019, Roh, Kim et al. 2020, Lin, Chen et al. 2021, Wu, Zhong et al. 2022, Liu, Zhang et al. 2024). Additionally, Peak-Alpha Frequency (PAF) (Arns, Gordon et al. 2015) and cordance (Cook, Hunter et al. 2014) were compared between patients with depression and healthy participants in one study, respectively.

##### Semiquantitative Analyses

Figure 3 shows the results of group comparisons for band-specific power and alpha asymmetry.

**Figure 3:**
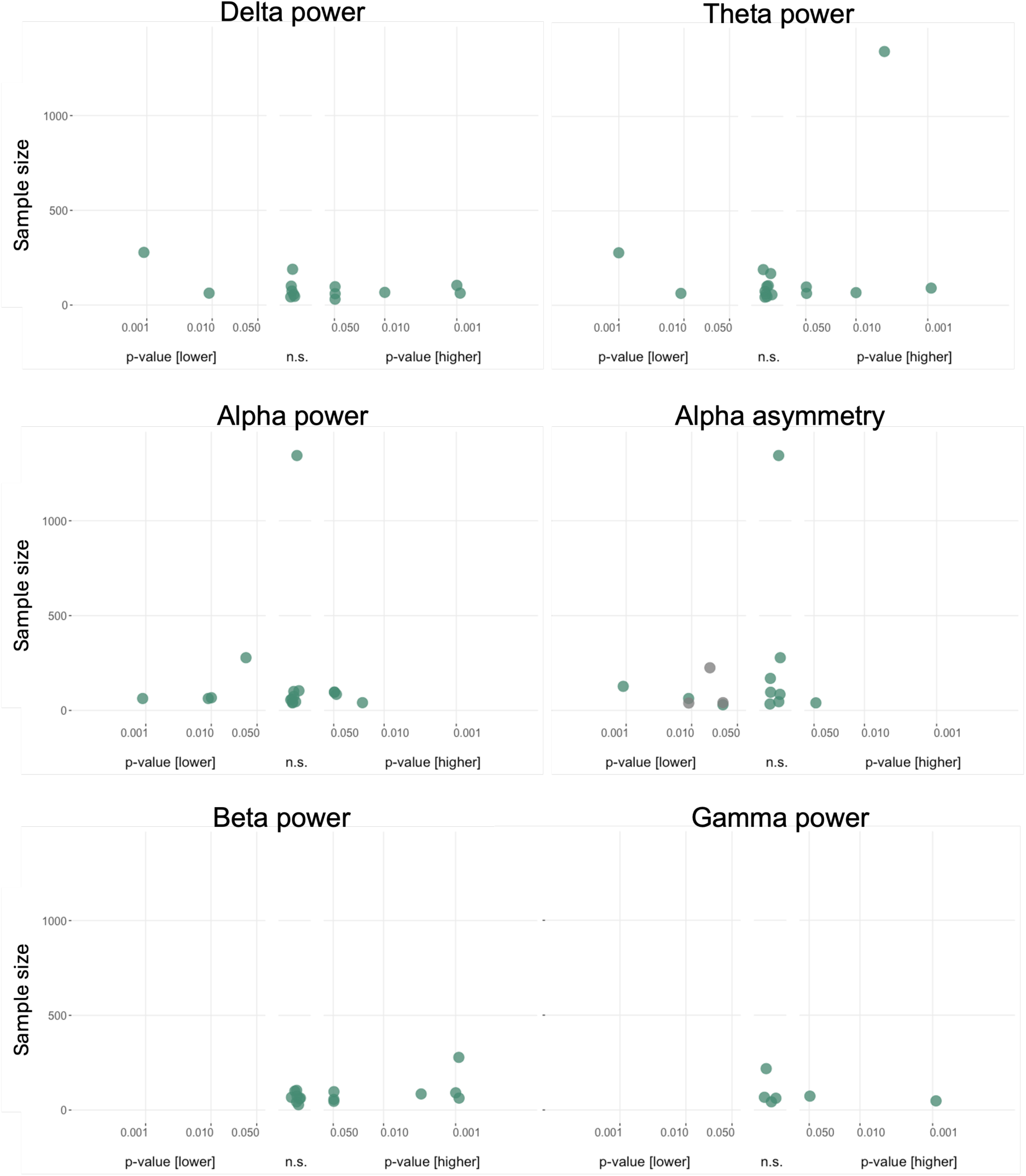
Albatross plots of group comparisons in band-specific power and alpha asymmetry. Power differences for group comparisons between patients and healthy participants. P values on the x-axis are displayed on a logarithmic scale (log10). Higher values in patients compared to healthy participants are depicted on the right-hand side, non-significant differences in the middle and lower values on the left-hand side of each panel. The total sample size for single studies is depicted on the y-axis. n.s., not significant. Due to the adaptation of directionality in alpha asymmetry (right-left), lower values indicate a predominance of left-sided frontal alpha activity.

*Power* in the *delta band* was compared by 13 studies. Six studies reported higher (Morgan, Witte et al. 2005, Begić, Popović-Knapić et al. 2011, Čukić, Stokić et al. 2020, Das and Yadav 2020, Huang, Yi et al. 2023, Xia, Wang et al. 2023), five reported no difference (Knott, Mahoney et al. 2001, Korb, Cook et al. 2008, Liu, Liu et al. 2022, Liu, Liu et al. 2022, Jiang, Huang et al. 2023), and two had lower (Mumtaz, Xia et al. 2017, Lin, Chen et al. 2021) values for patients than healthy participants. *Theta band* power was compared in 16 studies. Five found higher values (Begić, Popović-Knapić et al. 2011, Arns, Etkin et al. 2015, Huang, Yi et al. 2023, Lin, Du et al. 2023, Xia, Wang et al. 2023), nine no differences (Knott, Mahoney et al. 2001, Morgan, Witte et al. 2005, Korb, Cook et al. 2008, Cook, Hunter et al. 2014, Das and Yadav 2020, Hill, Zomorrodi et al. 2021, Liu, Liu et al. 2022, Liu, Liu et al. 2022, Jiang, Huang et al. 2023), and two studies found lower patient values (Mumtaz, Xia et al. 2017, Lin, Chen et al. 2021). Seventeen studies reported comparisons of *alpha power.* Four reported higher values (Dharmadhikari, Jaiswal et al., Jaworska, Blier et al. 2012, Wu, Zhong et al. 2022, Xia, Wang et al. 2023), nine reported no difference (Knott, Mahoney et al. 2001, Morgan, Witte et al. 2005, Korb, Cook et al. 2008, Arns, Etkin et al. 2015, Mumtaz, Xia et al. 2017, Das and Yadav 2020, Hill, Zomorrodi et al. 2021, Liu, Liu et al. 2022, Liu, Liu et al. 2022), and four reported lower values (Begić, Popović-Knapić et al. 2011, Mumtaz, Xia et al. 2017, Lin, Chen et al. 2021, Huang, Yi et al. 2023) in patients. *Beta power* was assessed in 15 studies. Seven studies found higher values (Knott, Mahoney et al. 2001, Begić, Popović-Knapić et al. 2011, Lin, Chen et al. 2021, Liu, Liu et al. 2022, Wu, Zhong et al. 2022, Lin, Du et al. 2023, Xia, Wang et al. 2023), eight did not find a difference (Morgan, Witte et al. 2005, Korb, Cook et al. 2008, Plante, Goldstein et al. 2013, Mumtaz, Xia et al. 2017, Das and Yadav 2020, Hill, Zomorrodi et al. 2021, Liu, Liu et al. 2022, Huang, Yi et al. 2023), and none found lower values in patients compared to healthy participants. Six studies reported comparisons in *gamma power,* with two studies showing higher values (Strelets, Garakh et al. 2007, Zeng, Lao et al. 2024), four studies showing no difference (Arikan, Gunver et al. 2019, Hill, Zomorrodi et al. 2021, Huang, Yi et al. 2023, Jang, Kim et al. 2023), and none showing lower values.

*Alpha asymmetry* was compared between patients and healthy controls in 14 studies, all of which reported results from frontal brain regions/electrodes. One study found higher values (Koo-Poeggel, Berger et al. 2019), seven found no difference (Knott, Mahoney et al. 2001, Segrave, Cooper et al. 2011, Jaworska, Blier et al. 2012, Arns, Bruder et al. 2016, Lin, Chen et al. 2021, Wu, Zhong et al. 2022, Liu, Zhang et al. 2024), and six found lower values in patients than healthy participants (Dharmadhikari, Jaiswal et al., Allen and Cohen 2010, Kemp, Griffiths et al. 2010, Cantisani, Koenig et al. 2015, Mumtaz, Xia et al. 2017, Roh, Kim et al. 2020).

Thus, semiquantitative analyses indicate an increase in low (delta and theta) and high (beta and gamma) frequency band power as well as alpha asymmetry with a relative increase in left-sided oscillatory activity in patients compared to healthy participants.

##### Meta-Analysis

If four or more studies provided sufficient information, a meta-analysis was performed. Figure 4 shows the results for band-specific power and alpha asymmetry. Meta-analysis was not possible for *gamma* power since sufficient information was not available.

**Figure 4:**
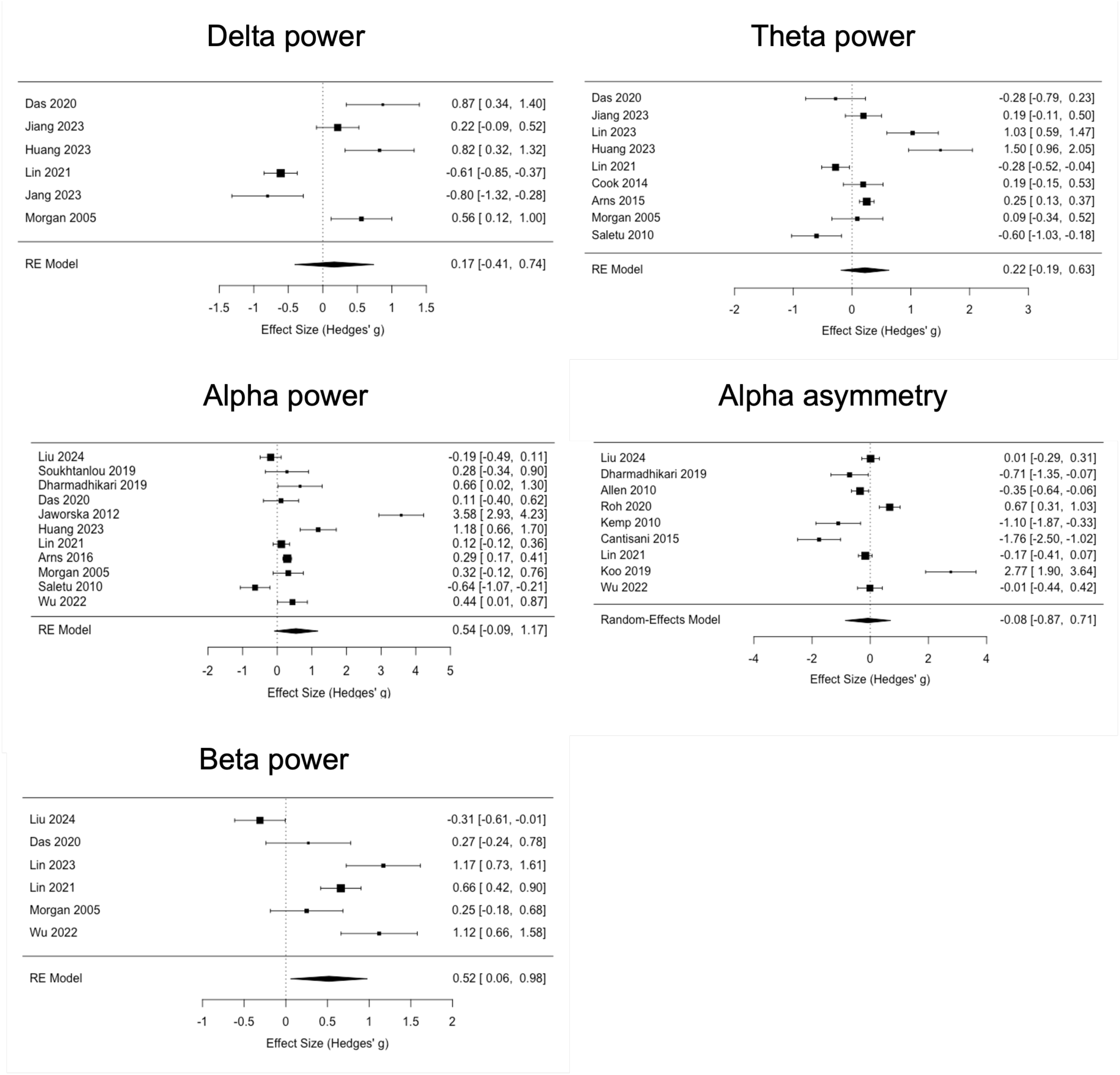
Forest plots of meta-analysis for comparisons of band-specific power and alpha asymmetry. RE = random effects. Due to the adaptation of directionality in alpha asymmetry (right-left), lower values indicate a predominance of left-sided frontal alpha activity.

*Band-specific power* comparison in the *delta* band yielded non-significant results with a high degree of heterogeneity among studies (k=6 studies, *Hedges’ g* = *0.17*, *95% CI -0.41-0.74*; heterogeneity: *I^2^* = *92.3%*, *p [Q]* < *0.001*). A similar picture was obtained for *theta* (k=9 studies, *Hedges’ g* = *0.22*, *95% CI -0.19-0.63*; heterogeneity: *I^2^* = *93.7%*, *p [Q]* < *0.001*) and alpha power (k=12 studies, *Hedges’ g* = *0.50*, *95% CI -0.07-1.08*; heterogeneity: *I^2^* = *97.3%*, *p [Q]* < *0.001*). For *beta power,* a significant group difference was obtained, with higher values in patients compared to healthy participants (k=6 studies, *Hedges’ g* = *0.52*, *95% CI -0.06-0.98*; heterogeneity: *I^2^* = *88.9%*, *p [Q]* < *0.001*) but considerable heterogeneity.

*Alpha asymmetry* did not differ between groups (k=9, *Hedges’ g* = -*0.08*, *95% CI -0.87-0.71*; heterogeneity: *I^2^* = *81.4%*, *p [Q]* < *0.001*), and heterogeneity was high.

Funnel plots suggested some asymmetry, especially for alpha power, but Egger’s test did not show evidence of publication bias (p>0.5) in all group comparisons (Supplementary Figure S2).

In summary, a meta-analysis confirmed an increase in beta power in patients with depression compared to healthy participants.

##### Narrative synthesis

Regarding group comparison of other EEG parameters, one study reported that PAF did not differ between the two groups (Arns, Gordon et al. 2015). Moreover, one study found that *cordance* was higher in patients than in healthy participants (Cook, Hunter et al. 2014), especially in the theta band.

#### Correlations with disease severity

##### Semiquantitative Analyses

Figure 5 summarizes the results of the correlation analysis of band-specific power and alpha asymmetry with disease severity. The number of studies available for *gamma* power was insufficient for semiquantitative analysis.

**Figure 5:**
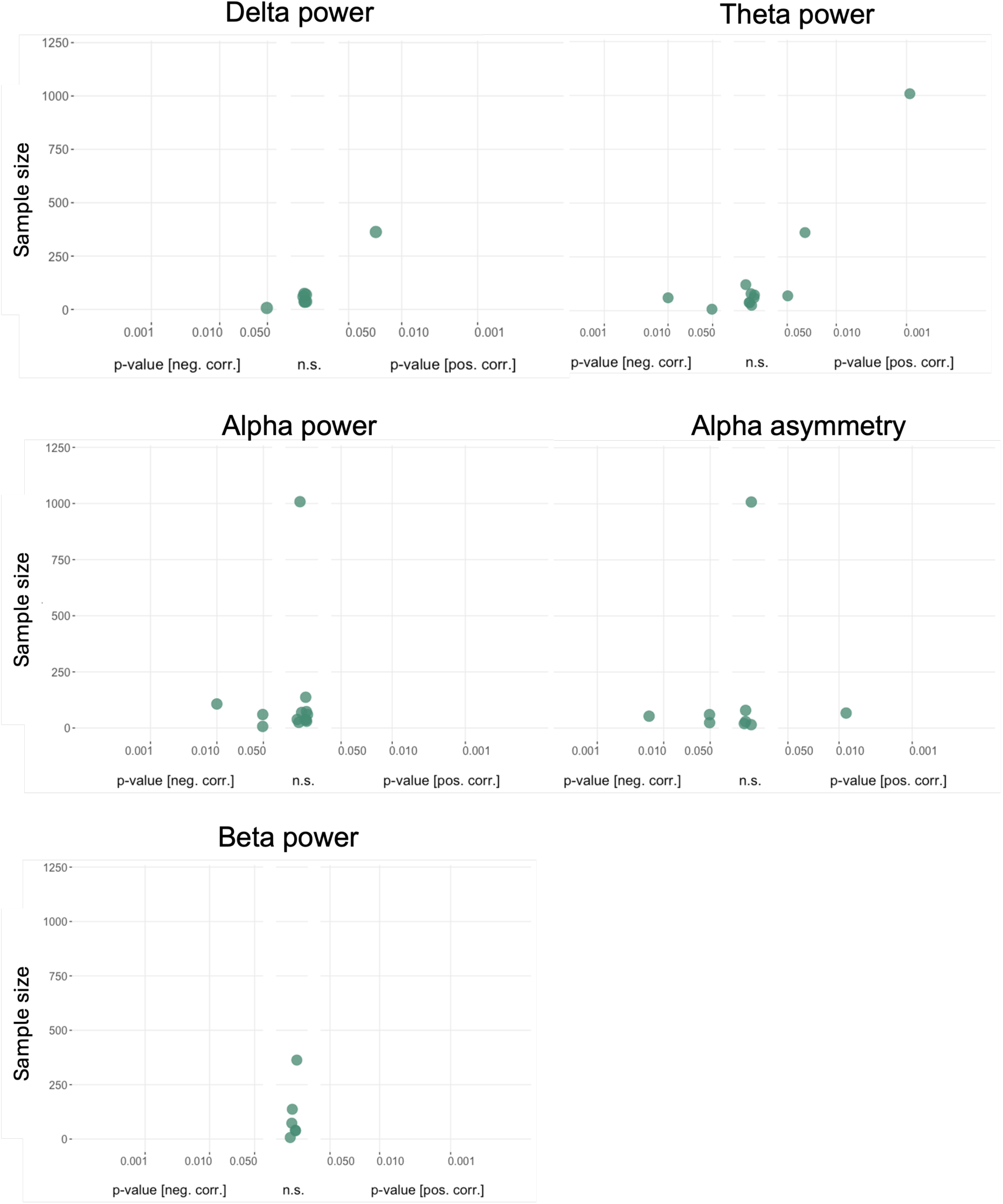
Albatross plots for band-specific power and alpha asymmetry correlations with disease severity. Plots show correlations of corresponding parameters with disease severity. P values of correlations are displayed on the x-axis on a logarithmic scale (log10). Positive correlations are depicted on the right-hand side, non-significant differences in the middle, and negative correlations on the left-hand side of each panel. The sample size for single studies is depicted on the y-axis. n.s., not significant.

Correlations between *band-specific power* in the *delta* range and depression severity were assessed in eight studies. One study showed a positive (Kesebir, Yosmaoglu et al. 2022), one study a negative (Farahbod, Cook et al. 2010), and six studies no significant correlation (Knott, Mahoney et al. 2001, Korb, Cook et al. 2008, Escolano, Navarro-Gil et al. 2014, Roh, Park et al. 2016, Kim, Oh et al. 2019, Huang, Yi et al. 2023). Twelve studies performed correlations for theta power. Three reported positive (Knott, Mahoney et al. 2001, Arns, Etkin et al. 2015, Kesebir, Yosmaoglu et al. 2022), two negative (Farahbod, Cook et al. 2010, Saletu, Anderer et al. 2010), and seven no significant correlation (Korb, Cook et al. 2008, Gold, Fachner et al. 2013, Cook, Hunter et al. 2014, Escolano, Navarro-Gil et al. 2014, Tas, Cebi et al. 2015, Roh, Park et al. 2016, Kim, Oh et al. 2019). Power in the *alpha* range was correlated with depression severity in 13 studies. No study found a positive correlation; three reported negative correlations (Farahbod, Cook et al. 2010, Saletu, Anderer et al. 2010, Zoon, Veth et al. 2013), and 10 reported non-significant correlations (Knott, Mahoney et al. 2001, Korb, Cook et al. 2008, Escolano, Navarro-Gil et al. 2014, Tas, Cebi et al. 2015, Arns, Bruder et al. 2016, Roh, Park et al. 2016, Kim, Oh et al. 2019, Das and Yadav 2020, Lin, Chen et al. 2021, Huang, Yi et al. 2023). Six studies performed correlations with *beta power*, and all reported non-significant results (Farahbod, Cook et al. 2010, Roh, Park et al. 2016, Kim, Oh et al. 2019, Lin, Chen et al. 2021, Kesebir, Yosmaoglu et al. 2022, Wu, Zhong et al. 2022).

Correlations between *alpha asymmetry* and depression severity were assessed in 9 studies, all reporting results from frontal brain regions/electrodes. Of those, one study reported a positive (Roh, Kim et al. 2020), three a negative (Saletu, Anderer et al. 2010, Jaworska, Blier et al. 2012, Marcu, Szekely-Copîndean et al. 2023), and five a non-significant relationship (Allen, Urry et al. 2004, Kemp, Griffiths et al. 2010, Gold, Fachner et al. 2013, Cantisani, Koenig et al. 2015, Arns, Bruder et al. 2016).

In summary, semiquantitative analyses do not point towards a consistent relationship between band-specific power and alpha asymmetry with depression severity.

##### Meta-Analysis

If four or more studies provided sufficient information, a meta-analysis was performed. Figure 6 shows the results for band-specific power and alpha asymmetry. Meta-analysis was not possible for *gamma* power since sufficient information was not available.

**Figure 6:**
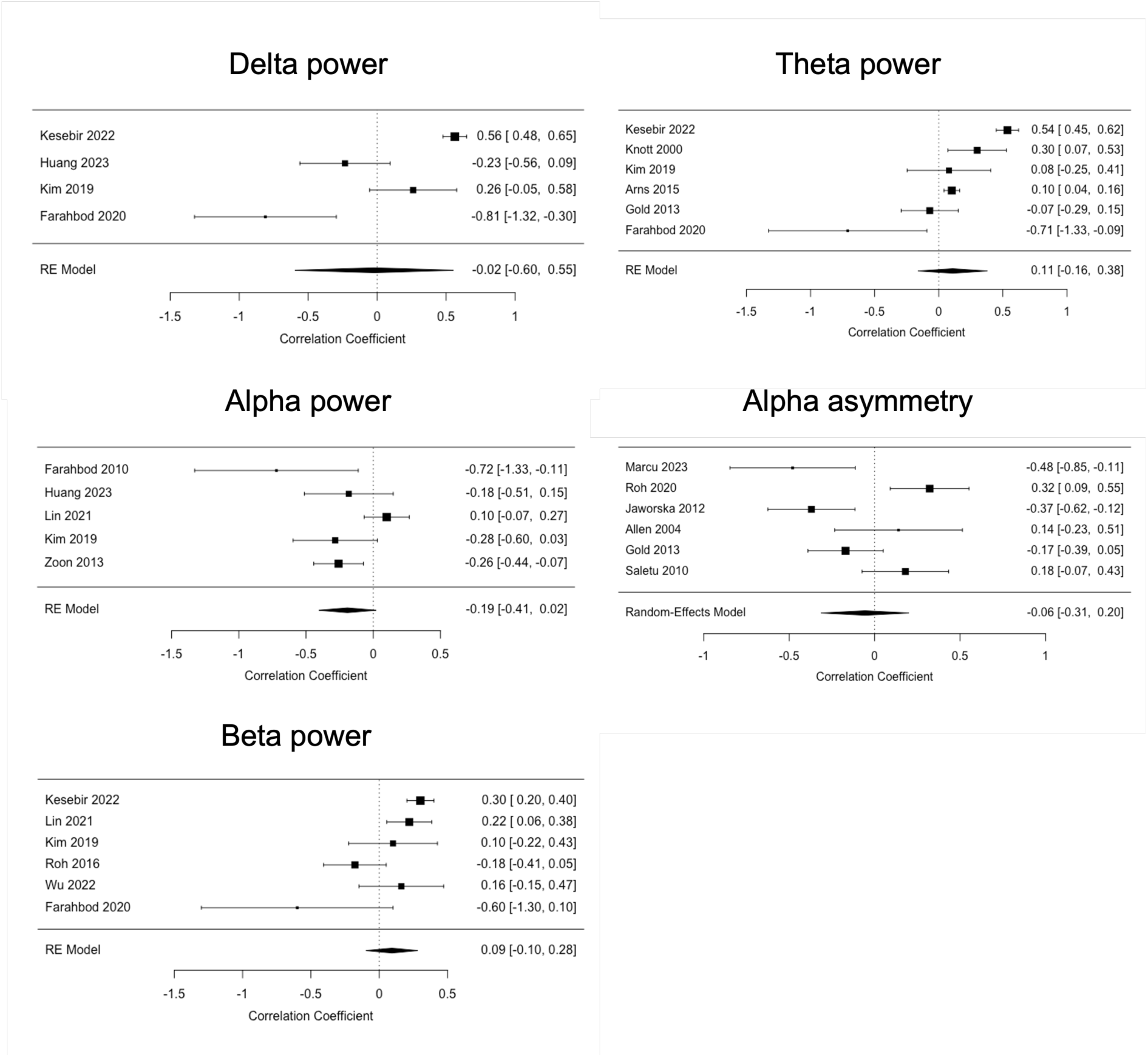
Forest plots of meta-analysis for correlations of band-specific power and alpha asymmetry with disease severity. RE = random effects.

Correlations between depression severity and *band-specific power* in the *delta* (k=4 studies, *r* = -*0.02*, *95% CI -0.60-0.55*; heterogeneity: *I^2^* = *94.3%*, *p [Q]* < *0.001*) and *theta* band (k=6 studies, *r* = *0.11*, *95% CI -0.16-0.38*; heterogeneity: *I^2^* = *94.8%*, *p [Q]* < *0.001*) were non-significant and showed very high heterogeneity. Correlations with *alpha* power were also non-significant and showed moderate heterogeneity (k=5 studies, *r* = *-0.19*, *95% CI -0.41-0.02*; heterogeneity: *I^2^* = *67.5%*, *p [Q]* < *0.01*). For *beta power,* a non-significant correlation was obtained, showing moderate heterogeneity (k=6, *r* = -*0.09*, *95% CI -0.10-0.28*; heterogeneity: *I^2^* = *75.4%*, *p [Q]* < *0.01*).

*Alpha asymmetry* was not correlated with depression severity (k=6, *r* = -*0.06*, *95% CI -0.31-0.20*; heterogeneity:*I^2^* = *81,4%*, *p [Q]* < *0.001*), and heterogeneity was high. Funnel plots suggested some asymmetry, especially for delta and beta power. However, Egger’s test did not show evidence of publication bias (p>0.5) in the reported correlations (Supplementary Figure S3).

Taken together, no consistent relationship of band-specific power and alpha asymmetry with depression severity was found in the semiquantitative and meta-analyses.

##### Narrative synthesis

*PAF* was related to depression severity in two studies. One study reported a significant positive correlation (Zhou, Wu et al. 2023) and another no relationship (Arns, Gordon et al. 2015).

Correlations of *cordance* with depression severity were performed in two studies. One study reported significant correlations of *cordance* in the alpha band with depression severity but with opposing directionality for different brain regions and non-significant correlations in the theta band (Scanlon, Jain et al. 2016). The other study reported no significant correlation between *cordance* and depression severity (Cook, Hunter et al. 2014).

## Discussion

The present study found evidence for an increase in the power of beta-oscillations and inconclusive evidence for an increase in slow frequency oscillations in the delta and theta range, as well as alpha asymmetry with relatively increased left-sided frontal oscillatory activity in patients with depression compared to healthy participants.

### Oscillatory brain activity in depression

The semiquantitative and meta-analytic results presented here in principle align with findings from previous narrative reviews reporting an increase in delta, theta, and beta oscillations (Newson and Thiagarajan 2019) as well as a potential role for slow (mainly theta) and high (primarily gamma) frequency oscillations and a recent meta-analysis pointing towards left frontal alpha asymmetry as potential diagnostic biomarkers in depression (de Aguiar Neto and Rosa 2019, Tsai, Li et al. 2023, Luo, Tang et al. 2025). *Beta oscillations* have been implicated in the top-down control of cortico-limbic circuits in depression (Hoy, de Hemptinne et al. 2023, Amemori, Graybiel et al. 2024, Xiao, Adkinson et al. 2024). They appear to be specifically involved in reward learning and reward biases in patients with depression and have been discussed as a potential biomarker of anhedonia (Xiao, Adkinson et al. 2024). Furthermore, experimental evidence shows that *beta oscillations* determine effort-related aspects and *theta oscillations* reward-related aspects of reward learning in depression. Additionally, increased *theta oscillations* were associated with negative experiences in patients with depression (Riddle, Alexander et al. 2020) and differentiated patients with MDD from those with anxiety disorder (Zhang, Lei et al. 2022). The role of theta oscillations in anxiety, as the most prominent comorbidity of depression, remains to be elucidated and warrants further study (Newson and Thiagarajan 2019). *Theta oscillations* have also been previously discussed as potential monitoring biomarkers in depression, e.g., predicting therapeutic response for non-invasive brain stimulation (Bailey, Hoy et al. 2018), and thus as a treatment target (Tsai, Li et al. 2023). However, the present study does not find a relationship between brain oscillations at any frequency and disease severity that would suggest potential as a monitoring biomarker. This does not mean that such a relationship does not exist, but that the current systematic review and meta-analysis of existing evidence cannot detect it.

*Alpha asymmetry* in depression has been conceptually related to an imbalance in appetitive and aversive motivational processing, which may correspond to negative and positive affect (van der Vinne, Vollebregt et al. 2017). A widely described predominance of left-sided frontal alpha activity was found to be correlated with sensitivity of the behavioral activation system (i.e., a deficiency in motivation) and hypothesized to be associated with anhedonia in previous literature (van der Vinne, Vollebregt et al. 2017). Our inconclusive findings regarding left alpha asymmetry reflect the prior literature, including recent systematic reviews and meta-analyses, with some showing such an effect (Luo, Tang et al. 2025) and others not (van der Vinne, Vollebregt et al. 2017). Both of these previous meta-analyses critically discuss the specificity and the role of methodological heterogeneity in left frontal alpha asymmetry, thereby questioning its use as a (single) diagnostic biomarker for depression.

Thus, our data point towards a potential role for beta oscillations and, to a much lesser extent, also theta oscillations and left frontal alpha asymmetry as potential diagnostic but not monitoring biomarkers in depression.

### Potential transdiagnostic implications

Similar to the present observations in patients with depression, combinations of increases in slow (especially theta) and high (especially beta) frequencies have been described in patients with chronic pain and pathological fatigue, thereby raising the question of specificity (Heitmann, Zebhauser et al. 2023, Zebhauser, Hohn et al. 2023, Zebhauser, Heitmann et al. 2024). These symptoms are highly comorbid and overlap in their anhedonic valence, which has fostered a discussion of reward deficiency as a potential common underlying pathological mechanism (Heitmann, Andlauer et al. 2020). A common electrophysiological model explaining this comorbidity is thalamocortical dysrhythmia. This model proposes that abnormal thalamocortical theta oscillations cause alterations in higher frequency bands in the beta and gamma range, resulting in different neuropsychiatric symptoms, including pain and depression (Llinas, Ribary et al. 1999, Sarnthein, Morel et al. 2005, Schulman, Cancro et al. 2011, De Ridder and Vanneste 2024). Recently, this model was also applied to neuropsychiatric disorders associated with reward deficiency and dopaminergic dysfunction, highlighting a role for theta and beta oscillations in the anterior cingulate cortex (ACC) and ventromedial prefrontal cortex (vmPFC) in these conditions (De Ridder and Vanneste 2024). This further supports the notion that aberrant theta and beta oscillations might reflect a common transdiagnostic pathomechanism related to altered reward processing and dopaminergic dysfunction across various disorders.

### Risk of bias and limitations

Different factors limit the interpretation and generalizability of the present results. They are related to the included studies and the applied analyses.

First, there was a considerable RoB, primarily due to a lack of control for relevant comorbidity (especially anxiety), potential medication effects, and non-blinded EEG assessment. Second, the sample sizes of the studies were relatively low (median n=68). The effects reported were observed in group comparisons, where sample size calculation indicates that for detecting medium effect sizes with a power of 80% (two-tailed t-test, *alpha*=0.05, *1-beta*=0.8), a total sample size of n=128 would be appropriate (Faul, Erdfelder et al. 2007). Thus, most studies included were underpowered, which increases the risk of false negative and false positive findings (Button, Ioannidis et al. 2013). Third, there was considerable methodological heterogeneity, especially regarding outcome parameters. However, due to the relatively small number of studies, results had to be pooled irrespective of the methods applied, e.g., local and global power results, data from eyes open and eyes closed EEG measurements, and using different recording systems and electrode placements. This can obscure specific effects but render the results obtained despite this heterogeneity more robust. However, deciding to include only the strongest effect in the case of multiple results for different brain regions being reported introduces a potential selection bias for false positive results. Fourth, the number of included studies differed substantially for different EEG measures. Only half of the studies could be included in meta-analyses for some EEG parameters, introducing a potential bias. For example, semiquantitative analyses pointed toward increased delta and theta oscillations, but meta-analyses did not yield significant results. However, for delta oscillations, only 3/6 studies, and for theta, only 3/5 studies reporting higher power in patients could be included in the meta-analyses due to a lack of detailed information needed for the meta-analyses. Still, for both frequency bands, 2/2 studies reporting lower power were included. This was vice versa for alpha power. Here, only 1/4 studies from the semiquantitative analyses reporting lower, but 4/4 studies reporting higher power in patients could be included in the meta-analysis, which then suggested a trend towards higher alpha power. Fifth, the fact that beta power did not correlate with disease severity limits its potential applicability as a biomarker to diagnostic but not monitoring purposes. Sixth, the insights from this study focus on EEG power measures and do not cover other EEG parameters in depression, e.g., connectivity measures, 1/f non-oscillatory brain activity, and microstate analyses. However, as stated in the introduction, the focus on frequently analyzed EEG parameters was chosen to maximize the number of studies included, e.g., to allow for meta-analysis.

### Conclusions

The present systematic review and meta-analysis provide a rigorous overview of EEG power-based measures in depression, yielding evidence for high beta power and inconclusive evidence for increased delta and theta power and left frontal alpha asymmetry in patients with depression. This provides insights into the pathophysiology of depression. However, the high RoB of studies highlights the need for well-powered and standardized studies to further evaluate the potential of these parameters as biomarkers and treatment targets in depression. The present results might motivate future studies combining non-invasive brain stimulation techniques targeting low and high oscillatory brain activity, such as theta-burst stimulation (Kishi, Ikuta et al. 2024).

## Data Availability

All data produced in the present study are available upon reasonable request to the authors

## References

Allen, J. J. B. and M. X. Cohen (2010). "Deconstructing the “Resting” State: Exploring the Temporal Dynamics of Frontal Alpha Asymmetry as an Endophenotype for Depression." Frontiers in Human Neuroscience 4.

Allen, J. J. B., H. L. Urry, S. K. Hitt and J. A. Coan (2004). "The stability of resting frontal electroencephalographic asymmetry in depression." Psychophysiology 41(2): 269–280.

Amemori, S., A. M. Graybiel and K. I. Amemori (2024). "Cingulate microstimulation induces negative decision-making via reduced top-down influence on primate fronto-cingulo-striatal network." Nat Commun 15(1): 4201.

Arikan, M. K., M. G. Gunver, N. Tarhan and B. Metin (2019). "High-Gamma: A biological marker for suicide attempt in patients with depression." Journal of Affective Disorders 254: 1–6.

Arns, M., G. Bruder, U. Hegerl, C. Spooner, D. M. Palmer, A. Etkin, K. Fallahpour, J. M. Gatt, L. Hirshberg and E. Gordon (2016). "EEG alpha asymmetry as a gender-specific predictor of outcome to acute treatment with different antidepressant medications in the randomized iSPOT-D study." Clinical Neurophysiology 127(1): 509–519.

Arns, M., A. Etkin, U. Hegerl, L. M. Williams, C. DeBattista, D. M. Palmer, P. B. Fitzgerald, A. Harris, R. deBeuss and E. Gordon (2015). "Frontal and rostral anterior cingulate (rACC) theta EEG in depression: Implications for treatment outcome?" European Neuropsychopharmacology 25(8): 1190–1200.

Arns, M., E. Gordon and N. N. Boutros (2015). "EEG Abnormalities Are Associated With Poorer Depressive Symptom Outcomes With Escitalopram and Venlafaxine-XR, but Not Sertraline: Results From the Multicenter Randomized iSPOT-D Study." Clinical EEG and Neuroscience 48(1): 33–40.

Bailey, N. W., K. E. Hoy, N. C. Rogasch, R. H. Thomson, S. McQueen, D. Elliot, C. M. Sullivan, B. D. Fulcher, Z. J. Daskalakis and P. B. Fitzgerald (2018). "Responders to rTMS for depression show increased fronto-midline theta and theta connectivity compared to non-responders." Brain Stimul 11(1): 190–203.

Begić, D., V. Popović-Knapić, J. Grubisin, B. Kosanović Rajačić, I. Filipcic, I. Telarovic and M. Jakovljevic (2011). "Quantitative electroencephalography in schizophrenia and depression." Psychiatria Danubina 23: 355–362.

Button, K. S., J. P. Ioannidis, C. Mokrysz, B. A. Nosek, J. Flint, E. S. Robinson and M. R. Munafo (2013). "Power failure: why small sample size undermines the reliability of neuroscience." Nat Rev Neurosci 14(5): 365–376.

Cantisani, A., T. Koenig, H. Horn, T. Müller, W. Strik and S. Walther (2015). "Psychomotor retardation is linked to frontal alpha asymmetry in major depression." Journal of Affective Disorders 188: 167–172.

Collaborators, G. B. D. M. D. (2022). "Global, regional, and national burden of 12 mental disorders in 204 countries and territories, 1990-2019: a systematic analysis for the Global Burden of Disease Study 2019." Lancet Psychiatry 9(2): 137–150.

Cook, I. A., A. M. Hunter, A. S. Korb and A. F. Leuchter (2014). "Do prefrontal midline electrodes provide unique neurophysiologic information in Major Depressive Disorder?" Journal of Psychiatric Research 53: 69–75.

Covidence (2021). Covidence systematic review software. Melbourne, Australia., Veritas Health Innovation.

Čukić, M., M. Stokić, S. Radenković, M. Ljubisavljević, S. Simić and D. Savić (2020). "Nonlinear analysis of EEG complexity in episode and remission phase of recurrent depression." International Journal of Methods in Psychiatric Research 29(2): e1816.

Das, J. and S. Yadav (2020). "Resting State Quantitative Electroencephalogram Power Spectra in Patients with Depressive Disorder as Compared to Normal Controls: An Observational Study." Indian Journal of Psychological Medicine 42(1): 30–38.

de Aguiar Neto, F. S. and J. L. G. Rosa (2019). "Depression biomarkers using non-invasive EEG: A review." Neuroscience & Biobehavioral Reviews 105: 83–93.

de la Salle, S., N. Jaworska, P. Blier, D. Smith and V. Knott (2020). "Using prefrontal and midline right frontal EEG-derived theta cordance and depressive symptoms to predict the differential response or remission to antidepressant treatment in major depressive disorder." Psychiatry Research: Neuroimaging 302.

De Ridder, D. and S. Vanneste (2024). "Thalamocortical dysrhythmia and reward deficiency syndrome as uncertainty disorders." Neuroscience 563: 20–32.

Dharmadhikari, A. S., S. V. Jaiswal, A. L. Tandle, D. Sinha and N. Jog "Study of Frontal Alpha Asymmetry in Mild Depression: A Potential Biomarker or Not?" 10.

Escolano, C., M. Navarro-Gil, J. Garcia-Campayo, M. Congedo, D. De Ridder and J. Minguez (2014). "A controlled study on the cognitive effect of alpha neurofeedback training in patients with major depressive disorder." Frontiers in Behavioral Neuroscience 8.

Farahbod, H., I. A. Cook, A. S. Korb, A. M. Hunter and A. F. Leuchter (2010). "Amygdala Lateralization at Rest and during Viewing of Neutral Faces in Major Depressive Disorder Using Low-Resolution Brain Electromagnetic Tomography." Clinical EEG and Neuroscience 41(1): 19–23.

Faul, F., E. Erdfelder, A. G. Lang and A. Buchner (2007). "G*Power 3: a flexible statistical power analysis program for the social, behavioral, and biomedical sciences." Behav Res Methods 39(2): 175–191.

FDA-NIH Biomarker Working Group. (2016). "BEST (Biomarkers, EndpointS, and other Tools) Resource." from https://www.ncbi.nlm.nih.gov/books/NBK326791/.

Fritz, C. O., P. E. Morris and J. J. Richler (2012). "Effect size estimates: current use, calculations, and interpretation." J Exp Psychol Gen 141(1): 2–18.

Gold, C., J. Fachner and J. ErkkilÄ (2013). "Validity and reliability of electroencephalographic frontal alpha asymmetry and frontal midline theta as biomarkers for depression." Scandinavian Journal of Psychology 54(2): 118–126.

Heitmann, H., T. F. M. Andlauer, T. Korn, M. Muhlau, P. Henningsen, B. Hemmer and M. Ploner (2020). "Fatigue, depression, and pain in multiple sclerosis: How neuroinflammation translates into dysfunctional reward processing and anhedonic symptoms." Mult Scler: 1352458520972279.

Heitmann, H., P. T. Zebhauser, V. D. Hohn, P. Henningsen and M. Ploner (2023). "Resting-state EEG and MEG biomarkers of pathological fatigue - A transdiagnostic systematic review." Neuroimage Clin 39: 103500.

Hill, A. T., R. Zomorrodi, I. Hadas, F. Farzan, D. Voineskos, A. Throop, P. B. Fitzgerald, D. M. Blumberger and Z. J. Daskalakis (2021). "Resting-state electroencephalographic functional network alterations in major depressive disorder following magnetic seizure therapy." Progress in Neuro-Psychopharmacology and Biological Psychiatry 108: 110082.

Hoy, C. W., C. de Hemptinne, S. S. Wang, C. J. Harmer, M. A. J. Apps, M. Husain, P. A. Starr and S. Little (2023). "Beta and theta oscillations track effort and previous reward in human basal ganglia and prefrontal cortex during decision making." bioRxiv.

Huang, Y., Y. Yi, Q. Chen, H. Li, S. Feng, S. Zhou, Z. Zhang, C. Liu, J. Li, Q. Lu, L. Zhang, W. Han, F. Wu and Y. Ning (2023). "Analysis of EEG features and study of automatic classification in first-episode and drug-naïve patients with major depressive disorder." BMC Psychiatry 23(1): 832.

Jackson, D. and R. Turner (2017). "Power analysis for random-effects meta-analysis." Res Synth Methods 8(3): 290–302.

Jang, K.-I., S. Kim, J.-H. Chae and C. Lee (2023). "Machine learning-based classification using electroencephalographic multi-paradigms between drug-naïve patients with depression and healthy controls." Journal of Affective Disorders 338: 270–277.

Jaworska, N., P. Blier, W. Fusee and V. Knott (2012). "Alpha power, alpha asymmetry and anterior cingulate cortex activity in depressed males and females." Journal of Psychiatric Research 46(11): 1483–1491.

Jiang, C., Z. Huang, Z. Zhou, L. Chen and H. Zhou (2023). "Decreased beta 1 (12–15 Hertz) power modulates the transfer of suicidal ideation to suicide in major depressive disorder." Acta Neuropsychiatrica 35(6): 362–371.

Kemp, A. H., K. Griffiths, K. L. Felmingham, S. A. Shankman, W. Drinkenburg, M. Arns, C. R. Clark and R. A. Bryant (2010). "Disorder specificity despite comorbidity: Resting EEG alpha asymmetry in major depressive disorder and post-traumatic stress disorder." Biological Psychology 85(2): 350–354.

Kesebir, S., A. Yosmaoglu and N. Tarhan (2022). "A dimensional approach to affective disorder: The relations between Scl-90 subdimensions and QEEG parameters." Frontiers in Psychiatry 13.

Kim, J. S., S. Oh, H. J. Jeon, K. S. Hong and J. H. Baek (2019). "Resting-state alpha and gamma activity in affective disorder with ADHD symptoms: Comparison between bipolar disorder and major depressive disorder." International Journal of Psychophysiology 143: 57–63.

Kishi, T., T. Ikuta, K. Sakuma, M. Hatano, Y. Matsuda, J. Wilkening, R. Goya-Maldonado, M. Tik, N. R. Williams, S. Kito and N. Iwata (2024). "Theta burst stimulation for depression: a systematic review and network and pairwise meta-analysis." Mol Psychiatry 29(12): 3893–3899.

Knott, V., C. Mahoney, S. Kennedy and K. Evans (2000). "Pre-Treatment EEG and It’s Relationship to Depression Severity and Paroxetine Treatment Outcome." Pharmacopsychiatry 33(06): 201–205.

Knott, V., C. Mahoney, S. Kennedy and K. Evans (2001). "EEG power, frequency, asymmetry and coherence in male depression." Psychiatry Research: Neuroimaging 106(2): 123–140.

Koo-Poeggel, P., C. Berger, G. Kronenberg, J. Bartz, P. Wybitul, O. Reis and J. Hoeppner (2019). "Combined cognitive, psychomotor and electrophysiological biomarkers in major depressive disorder." European Archives of Psychiatry and Clinical Neuroscience 269.

Korb, A. S., I. A. Cook, A. M. Hunter and A. F. Leuchter (2008). "Brain Electrical Source Differences between Depressed Subjects and Healthy Controls." Brain Topography 21(2): 138–146.

Leuchter, A. F. C., I.A.; Lufkin, R.B.; Dunkin, J.; Newton, T.F.; Cummings, J.L.; Mackey, J.K., Walter, D.O. (1994). "Cordance: a new method for assessment of cerebral perfusion and metabolism using quantitative electroencephalography." Neuroimage Jun;1(3): 208–219.

Lin, I. M., T.-C. Chen, H.-Y. Lin, S.-Y. Wang, J.-L. Sung and C.-W. Yen (2021). "Electroencephalogram patterns in patients comorbid with major depressive disorder and anxiety symptoms: Proposing a hypothesis based on hypercortical arousal and not frontal or parietal alpha asymmetry." Journal of Affective Disorders 282: 945–952.

Lin, S., Y. Du, Y. Xia, L. Xiao and G. Wang (2023). "Resting-state EEG as a potential indicator to predict sleep quality in depressive patients." International Journal of Psychophysiology 191: 1–8.

Liu, S., X. Liu, D. Yan, S. Chen, Y. Liu, X. Hao, W. Ou, Z. Huang, F. Su, F. He and D. Ming (2022). "Alterations in Patients With First-Episode Depression in the Eyes-Open and Eyes-Closed Conditions: A Resting-State EEG Study." IEEE Transactions on Neural Systems and Rehabilitation Engineering 30: 1019–1029.

Liu, X., S. Liu, M. Li, F. Su, S. Chen, Y. Ke and D. Ming (2022). "Altered gamma oscillations and beta–gamma coupling in drug-naive first-episode major depressive disorder: Association with sleep and cognitive disturbance." Journal of Affective Disorders 316: 99–108.

Liu, X., H. Zhang, Y. Cui, T. Zhao, B. Wang, X. Xie, S. Liang, S. Sha, Y. Yan, X. Zhao and L. Zhang (2024). "EEG-based major depressive disorder recognition by neural oscillation and asymmetry." Frontiers in Neuroscience 18.

Llinas, R. R., U. Ribary, D. Jeanmonod, E. Kronberg and P. P. Mitra (1999). "Thalamocortical dysrhythmia: A neurological and neuropsychiatric syndrome characterized by magnetoencephalography." Proc Natl Acad Sci U S A 96(26): 15222–15227.

Luo, Y., M. Tang and X. Fan (2025). "Meta analysis of resting frontal alpha asymmetry as a biomarker of depression." npj Mental Health Research 4(1).

Marcu, G. M., R. D. Szekely-Copîndean, A.-M. Radu, M. D. Bucuță, R. S. Fleacă, C. Tănăsescu, M. D. Roman, A. Boicean and C. I. Băcilă (2023). "Resting-state frontal, frontlateral, and parietal alpha asymmetry:A pilot study examining relations with depressive disorder type and severity." Frontiers in Psychology 14.

Marwaha, S., E. Palmer, T. Suppes, E. Cons, A. H. Young and R. Upthegrove (2023). "Novel and emerging treatments for major depression." Lancet 401(10371): 141–153.

Miljevic, A., N. W. Bailey, O. W. Murphy, M. P. N. Perera and P. B. Fitzgerald (2023). "Alterations in EEG functional connectivity in individuals with depression: A systematic review." J Affect Disord 328: 287–302.

Morgan, M., E. Witte, I. Cook, A. Leuchter, M. Abrams and B. Siegman (2005). "Influence of Age, Gender, Health Status, and Depression on Quantitative EEG." Neuropsychobiology 52: 71–76.

Muller, V. I., E. C. Cieslik, I. Serbanescu, A. R. Laird, P. T. Fox and S. B. Eickhoff (2017). "Altered Brain Activity in Unipolar Depression Revisited: Meta-analyses of Neuroimaging Studies." JAMA Psychiatry 74(1): 47–55.

Mumtaz, W., L. Xia, S. S. A. Ali, M. A. M. Yasin, M. Hussain and A. S. Malik (2017). "Electroencephalogram (EEG)-based computer-aided technique to diagnose major depressive disorder (MDD)." Biomedical Signal Processing and Control 31: 108–115.

Nemeroff, C. B. (2020). "The State of Our Understanding of the Pathophysiology and Optimal Treatment of Depression: Glass Half Full or Half Empty?" Am J Psychiatry 177(8): 671–685.

Newson, J. J. and T. C. Thiagarajan (2019). "EEG Frequency Bands in Psychiatric Disorders: A Review of Resting State Studies." Frontiers in Human Neuroscience 12.

Olbrich, S., R. van Dinteren and M. Arns (2015). "Personalized Medicine: Review and Perspectives of Promising Baseline EEG Biomarkers in Major Depressive Disorder and Attention Deficit Hyperactivity Disorder." Neuropsychobiology 72(3-4): 229–240.

Page, M. J., J. E. McKenzie, P. M. Bossuyt, I. Boutron, T. C. Hoffmann, C. D. Mulrow, L. Shamseer, J. M. Tetzlaff, E. A. Akl, S. E. Brennan, R. Chou, J. Glanville, J. M. Grimshaw, A. Hrobjartsson, M. M. Lalu, T. Li, E. W. Loder, E. Mayo-Wilson, S. McDonald, L. A. McGuinness, L. A. Stewart, J. Thomas, A. C. Tricco, V. A. Welch, P. Whiting and D. Moher (2021). "The PRISMA 2020 statement: an updated guideline for reporting systematic reviews." Syst Rev 10(1): 89.

Pizzagalli, D. A., J. B. Nitschke, T. R. Oakes, A. M. Hendrick, K. A. Horras, C. L. Larson, H. C. Abercrombie, S. M. Schaefer, J. V. Koger, R. M. Benca, R. D. Pascual-Marqui and R. J. Davidson (2002). "Brain electrical tomography in depression: the importance of symptom severity, anxiety, and melancholic features." Biological Psychiatry 52(2): 73–85.

Plante, D. T., M. R. Goldstein, E. C. Landsness, B. A. Riedner, J. J. Guokas, T. Wanger, G. Tononi and R. M. Benca (2013). "Altered overnight modulation of spontaneous waking EEG reflects altered sleep homeostasis in major depressive disorder: A high-density EEG investigation." Journal of Affective Disorders 150(3): 1167–1173.

Putnam, K. M. and L. B. McSweeney (2008). "Depressive symptoms and baseline prefrontal EEG alpha activity: A study utilizing Ecological Momentary Assessment." Biological Psychology 77(2): 237–240.

Quinn, C. R., C. J. Rennie, A. W. F. Harris and A. H. Kemp (2014). "The impact of melancholia versus non-melancholia on resting-state, EEG alpha asymmetry: Electrophysiological evidence for depression heterogeneity." Psychiatry Research 215(3): 614–617.

R Core Team (2021). R: A Language and Environement for Statistical Computing. Vienna, Austria, R Foundation for Statistical Computing.

Riddle, J., M. Alexander, T. McPherson, R. Lapate, C. Schiller, D. Rubinow and F. Frohlich (2020). "Theta Oscillations Increase During Negative Experiences in Patients in a Depressive Episode." Biological Psychiatry 87(9): S234–S235.

Roh, S.-C., J. S. Kim, S. Kim, Y. Kim and S.-H. Lee (2020). "Frontal Alpha Asymmetry Moderated by Suicidal Ideation in Patients with Major Depressive Disorder: A Comparison with Healthy Individuals." Clinical Psychopharmacology and Neuroscience 18(1): 58–66.

Roh, S.-C., E.-J. Park, M. Shim and S.-H. Lee (2016). "EEG beta and low gamma power correlates with inattention in patients with major depressive disorder." Journal of Affective Disorders 204: 124–130.

Saletu, B., P. Anderer and G. M. Saletu-Zyhlarz (2010). "EEG Topography and Tomography (LORETA) in Diagnosis and Pharmacotherapy of Depression." Clinical EEG and Neuroscience 41(4): 203–210.

Sarnthein, J., A. Morel, A. von Stein and D. Jeanmonod (2005). "Thalamocortical theta coherence in neurological patients at rest and during a working memory task." Int J Psychophysiol 57(2): 87–96.

Scanlon, G. C., F. A. Jain, A. M. Hunter, I. A. Cook and A. F. Leuchter (2016). "Neurophysiologic Correlates of Headache Pain in Subjects With Major Depressive Disorder." Clinical EEG and Neuroscience 48(3): 159–167.

Schulman, J. J., R. Cancro, S. Lowe, F. Lu, K. D. Walton and R. R. Llinas (2011). "Imaging of thalamocortical dysrhythmia in neuropsychiatry." Front Hum Neurosci 5: 69.

Segrave, R., N. Cooper, R. Thomson, R. Croft, D. Sheppard and P. Fitzgerald (2011). "Individualized Alpha Activity and Frontal Asymmetry in Major Depression." Clinical EEG and neuroscience: official journal of the EEG and Clinical Neuroscience Society (ENCS) 42: 45–52.

Shorey, S., E. D. Ng and C. H. J. Wong (2022). "Global prevalence of depression and elevated depressive symptoms among adolescents: A systematic review and meta-analysis." Br J Clin Psychol 61(2): 287–305.

Soukhtanlou, M., R. Rostami, M. A. Salehinejad, M. Gholamali Lavasani, A. Sharifi and A. Hekmatmanesh (2019). "Electrophysiological processing of happiness during conscious and sub-conscious awareness in depression." Neurology, Psychiatry and Brain Research 33: 32–38.

Strelets, V., Z. Garakh and V. Novototskii-Vlasov (2007). "Comparative study of the gamma rhythm in normal conditions, during examination stress, and in patients with first depressive episode." Neuroscience and behavioral physiology 37: 387–394.

Tas, C., M. Cebi, O. Tan, G. Hızlı-Sayar, N. Tarhan and E. C. Brown (2015). "EEG power, cordance and coherence differences between unipolar and bipolar depression." Journal of Affective Disorders 172: 184–190.

Tsai, Y. C., C. T. Li and C. H. Juan (2023). "A review of critical brain oscillations in depression and the efficacy of transcranial magnetic stimulation treatment." Front Psychiatry 14: 1073984.

van der Vinne, N., M. A. Vollebregt, M. J. A. M. van Putten and M. Arns (2017). "Frontal alpha asymmetry as a diagnostic marker in depression: Fact or fiction? A meta-analysis." NeuroImage: Clinical 16: 79–87.

Viechtbauer, W. (2010). "Conducting Meta-Analyses in R with the metafor Package." Journal of Statistical Software 36(3): 1–48.

Voetterl, H., A. T. Sack, S. Olbrich, S. Stuiver, R. Rouwhorst, A. Prentice, D. A. Pizzagalli, N. van der Vinne, J. A. van Waarde, M. Brunovsky, I. van Oostrom, B. Reitsma, J. Fekkes, H. van Dijk and M. Arns (2023). "Alpha peak frequency-based Brainmarker-I as a method to stratify to pharmacotherapy and brain stimulation treatments in depression." Nature Mental Health 1(12): 1023–1032.

Wu, Z., X. Zhong, G. Lin, Q. Peng, M. Zhang, H. Zhou, Q. Wang, B. Chen and Y. Ning (2022). "Resting-state electroencephalography of neural oscillation and functional connectivity patterns in late-life depression." Journal of Affective Disorders 316: 169–176.

Xia, Y., G. Wang, L. Xiao, Y. Du, S. Lin, C. Nan and S. Weng (2023). "Effects of Early Adverse Life Events on Depression and Cognitive Performance from the Perspective of the Heart-Brain Axis." Brain Sciences 13: 1174.

Xiao, J., J. A. Adkinson, J. Myers, A. B. Allawala, R. K. Mathura, V. Pirtle, R. Najera, N. R. Provenza, E. Bartoli, A. J. Watrous, D. Oswalt, R. Gadot, A. Anand, B. Shofty, S. J. Mathew, W. K. Goodman, N. Pouratian, X. Pitkow, K. R. Bijanki, B. Hayden and S. A. Sheth (2024). "Beta activity in human anterior cingulate cortex mediates reward biases." Nat Commun 15(1): 5528.

Zebhauser, P. T., H. Heitmann, E. S. May and M. Ploner (2024). "Resting-state electroencephalography and magnetoencephalography in migraine-a systematic review and meta-analysis." J Headache Pain 25(1): 147.

Zebhauser, P. T., V. D. Hohn and M. Ploner (2023). "Resting-state electroencephalography and magnetoencephalography as biomarkers of chronic pain: a systematic review." Pain 164(6): 1200–1221.

Zeng, L. L., H. Shen, L. Liu, L. Wang, B. Li, P. Fang, Z. Zhou, Y. Li and D. Hu (2012). "Identifying major depression using whole-brain functional connectivity: a multivariate pattern analysis." Brain 135(Pt 5): 1498–1507.

Zeng, Y., J. Lao, Z. Wu, G. Lin, Q. Wang, M. Yang, S. Zhang, D. Xu, M. Zhang, S. Liang, Q. Liu, K. Yao, J. Li, Y. Ning and X. Zhong (2024). "Altered resting-state brain oscillation and the associated cognitive impairments in late-life depression with different depressive severity: An EEG power spectrum and functional connectivity study." Journal of Affective Disorders 348: 124–134.

Zhang, Y., L. Lei, Z. Liu, M. Gao, Z. Liu, N. Sun, C. Yang, A. Zhang, Y. Wang and K. Zhang (2022). "Theta oscillations: A rhythm difference comparison between major depressive disorder and anxiety disorder." Front Psychiatry 13: 827536.

Zhou, P., Q. Wu, L. Zhan, Z. Guo, C. Wang, S. Wang, Q. Yang, J. Lin, F. Zhang, L. Liu, D. Lin, W. Fu and X. Wu (2023). "Alpha peak activity in resting-state EEG is associated with depressive score." Frontiers in Neuroscience 17: 1057908.

Zoon, H. F. A., C. P. M. Veth, M. Arns, W. H. I. M. Drinkenburg, W. Talloen, P. J. Peeters and J. L. Kenemans (2013). "EEG Alpha Power as an Intermediate Measure Between Brain-Derived Neurotrophic Factor Val66Met and Depression Severity in Patients With Major Depressive Disorder." Journal of Clinical Neurophysiology 30(3).

