## Supplementary Material for "Resting-state EEG activity as a Biomarker and Treatment Target in Depression: A Systematic Review and Meta-analysis"

### Search Strategy:

Pubmed:

("depress*"[Title/Abstract] OR "depression"[MeSH Terms]) AND ("quantitative"[Title/Abstract] OR "spontaneous"[Title/Abstract] OR "rest*"[Title/Abstract]) AND ("qEEG"[Title/Abstract] OR "eeg"[Title/Abstract] OR "electroencephalogra*"[Title/Abstract] OR "electroencephalography"[MeSH Terms])

WoS:
("depress*") AND ("quantitative" OR "spontaneous" OR "rest*") AND ("qEEG" OR "eeg" OR "electroencephalogra*")

Embase:
(depress$.ab. or depress$.ti.) and (quantitative.ab. or quantitative.ti. or spontaneous.ab. or spontaneous.ti. or rest$.ab. or rest$.ti. or waking.ab. or waking.ti.) and (qEEG.ab. or qEEG.ti. or electroencephalogra$.ab. or electroencephalogra$.ti. or meg.ab. or meg.ti. or magnetoencephalogra$.ab. or magnetoencephalogra$.ti.)

### Supplementary Figure S1:

Risk of bias assessment:

| **Study ID** | **Title** | **Selection: Case definition** | **Selection: Representativeness** | **Selection: Controls** | **Selection: Definition of Controls** | **Comparability: Anxiety** | **Comparability: Any other factor** | **Outcome: Independent EEG preprocessing /analysis** | **Outcome: Description of EEG measurement** | **Outcome: Statistical test** |
|---|---|---|---|---|---|---|---|---|---|---|
| Allen 2004 | The stability of resting frontal electroencephalographic asymmetry in depression. | Low RoB | High RoB | N/a | N/a | High RoB | High RoB | High RoB | Low RoB | High RoB |
| Allen 2010 | Deconstructing the "resting" state: exploring the temporal dynamics of frontal alpha asymmetry as an endophenotype for depression. | Low RoB | High RoB | High RoB | High RoB | High RoB | High RoB | High RoB | Low RoB | High RoB |
| Arikan 2019 | High-Gamma: A biological marker for suicide attempt in patients with depression. | Low RoB | Low RoB | High RoB | High RoB | High RoB | High RoB | High RoB | Low RoB | Low RoB |
| Arns 2015 | Frontal and rostral anterior cingulate (rACC) theta EEG in depression: implications for treatment outcome? | Low RoB | Low RoB | Low RoB | Low RoB | High RoB | Low RoB | Low RoB | Low RoB | Low RoB |
| Arns 2016 | EEG alpha asymmetry as a gender-specific predictor of outcome to acute treatment with different antidepressant medications in the randomized iSPOT-D study. | Low RoB | Low RoB | High RoB | High RoB | High RoB | Low RoB | Low RoB | Low RoB | Low RoB |
| Arns 2017 | EEG Abnormalities Are Associated With Poorer Depressive Symptom Outcomes With Escitalopram and Venlafaxine-XR, but Not Sertraline: Results From the Multicenter Randomized iSPOT-D Study. | Low RoB | Low RoB | Low RoB | High RoB | Low RoB | Low RoB | Low RoB | Low RoB | Low RoB |
| Begić 2011 | Quantitative electroencephalography in schizophrenia and depression. | Low RoB | Low RoB | High RoB | Low RoB | High RoB | High RoB | High RoB | High RoB | High RoB |
| Cantisani 2015 | Psychomotor retardation is linked to frontal alpha asymmetry in major depression. | Low RoB | Low RoB | Low RoB | Low RoB | High RoB | Low RoB | High RoB | Low RoB | Low RoB |
| Cook 2014 | Do prefrontal midline electrodes provide unique neurophysiologic information in Major Depressive Disorder? | Low RoB | High RoB | Low RoB | Low RoB | High RoB | Low RoB | High RoB | Low RoB | Low RoB |
| Čukić 2020 | Nonlinear analysis of EEG complexity in episode and remission phase of recurrent depression. | Low RoB | High RoB | Low RoB | Low RoB | High RoB | High RoB | High RoB | Low RoB | High RoB |

| | | | | | | | | | | |
|---|---|---|---|---|---|---|---|---|---|---|
| Das 2020 | Resting State Quantitative Electroencephalogram Power Spectra in Patients with Depressive Disorder as Compared to Normal Controls: An Observational Study. | Low RoB | Low RoB | Low RoB | Low RoB | High RoB | High RoB | High RoB | Low RoB | High RoB |
| Dharmadhikari 2019 | Study of Frontal Alpha Asymmetry in Mild Depression: A Potential Biomarker or Not? | Low RoB | Low RoB | Low RoB | Low RoB | Low RoB | High RoB | High RoB | Low RoB | High RoB |
| Escolano 2014 | A controlled study on the cognitive effect of alpha neurofeedback training in patients with major depressive disorder. | Low RoB | Low RoB | N/a | N/a | High RoB | High RoB | High RoB | Low RoB | High RoB |
| Farahbod 2010 | Amygdala lateralization at rest and during viewing of neutral faces in major depressive disorder using low-resolution brain electromagnetic tomography. | Low RoB | Low RoB | N/a | N/a | High RoB | High RoB | High RoB | Low RoB | High RoB |
| Gold 2013 | Validity and reliability of electroencephalographic frontal alpha asymmetry and frontal midline theta as biomarkers for depression. | Low RoB | Low RoB | N/a | N/a | High RoB | High RoB | High RoB | Low RoB | High RoB |
| Hill 2021 | Resting-state electroencephalographic functional network alterations in major depressive disorder following magnetic seizure therapy. | Low RoB | Low RoB | High RoB | Low RoB | High RoB | High RoB | High RoB | Low RoB | High RoB |
| Huang 2023 | Analysis of EEG features and study of automatic classification in first-episode and drug-naïve patients with major depressive disorder. | Low RoB | Low RoB | High RoB | Low RoB | High RoB | High RoB | High RoB | Low RoB | Low RoB |
| Jang 2023 | Machine learning-based classification using electroencephalographic multi-paradigms between drug-naïve patients with depression and healthy controls. | Low RoB | Low RoB | High RoB | Low RoB | Low RoB | Low RoB | Low RoB | Low RoB | Low RoB |
| Jaworska 2012 | α Power, α asymmetry and anterior cingulate cortex activity in depressed males and females. | Low RoB | High RoB | High RoB | Low RoB | High RoB | Low RoB | High RoB | Low RoB | High RoB |
| Jiang 2023 | Decreased beta 1 (12-15 Hertz) power modulates the transfer of suicidal ideation to suicide in major depressive disorder. | Low RoB | Low RoB | Low RoB | Low RoB | High RoB | Low RoB | High RoB | Low RoB | Low RoB |
| Kemp 2010 | Disorder specificity despite comorbidity: resting EEG alpha asymmetry in major depressive disorder and post-traumatic stress disorder. | Low RoB | Low RoB | Low RoB | Low RoB | Low RoB | Low RoB | High RoB | Low RoB | High RoB |
| Kesebir 2022 | A dimensional approach to affective disorder: The relations between Scl-90 subdimensions and QEEG parameters | Low RoB | High RoB | N/a | N/a | High RoB | High RoB | High RoB | High RoB | Low RoB |
| Kim 2019 | Resting-state alpha and gamma activity in affective disorder with ADHD symptoms: Comparison between bipolar disorder and major depressive disorder. | Low RoB | High RoB | N/a | N/a | High RoB | Low RoB | High RoB | High RoB | High RoB |
| Knott 2000 | Pre-treatment EEG and it's relationship to depression severity and paroxetine treatment outcome. | Low RoB | High RoB | N/a | N/a | High RoB | High RoB | High RoB | High RoB | High RoB |
| Knott 2001 | EEG power, frequency, asymmetry and coherence in male depression. | Low RoB | High RoB | High RoB | Low RoB | High RoB | High RoB | High RoB | Low RoB | High RoB |
| Koo 2019 | Combined cognitive, psychomotor and electrophysiological biomarkers in major depressive disorder. | Low RoB | High RoB | Low RoB | Low RoB | High RoB | High RoB | High RoB | Low RoB | Low RoB |
| Korb 2008 | Brain electrical source differences between depressed subjects and healthy controls. | Low RoB | Low RoB | Low RoB | Low RoB | High RoB | Low RoB | Low RoB | Low RoB | High RoB |
| Lin 2021 | Electroencephalogram patterns in patients comorbid with major depressive disorder and anxiety symptoms: Proposing a hypothesis based on hypercortical arousal and not frontal or parietal alpha asymmetry. | Low RoB | High RoB | Low RoB | Low RoB | High RoB | High RoB | High RoB | High RoB | Low RoB |
| Lin 2023 | Resting-state EEG as a potential indicator to predict sleep quality in depressive patients. | Low RoB | High RoB | Low RoB | Low RoB | High RoB | Low RoB | High RoB | Low RoB | High RoB |
| Liu 2022 | Altered gamma oscillations and beta-gamma coupling in drug-naive first-episode major depressive disorder: Association with sleep and cognitive disturbance. | Low RoB | High RoB | Low RoB | Low RoB | High RoB | Low RoB | High RoB | Low RoB | High RoB |
| Liu 2022 | Alterations in Patients With First-Episode Depression in the Eyes-Open and Eyes-Closed Conditions: A Resting-State EEG Study. | Low RoB | High RoB | Low RoB | Low RoB | High RoB | Low RoB | High RoB | Low RoB | High RoB |
| Liu 2024 | EEG-based major depressive disorder recognition by neural oscillation and asymmetry. | Low RoB | Low RoB | High RoB | High RoB | High RoB | High RoB | High RoB | Low RoB | Low RoB |
| Marcu 2023 | Resting-state frontal, frontlateral, and parietal alpha asymmetry:A pilot study examining relations with depressive disorder type and severity. | Low RoB | Low RoB | N/a | N/a | Low RoB | Low RoB | High RoB | Low RoB | High RoB |
| Morgan 2005 | Influence of age, gender, health status, and depression on quantitative EEG. | Low RoB | High RoB | High RoB | High RoB | High RoB | Low RoB | Low RoB | Low RoB | High RoB |
| Mumtaz 2017 | Electroencephalogram (EEG)-based computer-aided technique to diagnose major depressive disorder (MDD) | Low RoB | High RoB | Low RoB | Low RoB | High RoB | High RoB | Low RoB | Low RoB | High RoB |
| Pizzagalli 2002 | Brain electrical tomography in depression: the importance of symptom severity, anxiety, and melancholic features. | Low RoB | Low RoB | Low RoB | Low RoB | Low RoB | Low RoB | High RoB | Low RoB | High RoB |
| Plante 2013 | Altered overnight modulation of spontaneous waking EEG reflects altered sleep homeostasis in major depressive disorder: a high-density EEG investigation. | Low RoB | Low RoB | Low RoB | Low RoB | High RoB | Low RoB | High RoB | Low RoB | High RoB |
| Putnam 2008 | Depressive symptoms and baseline prefrontal EEG alpha activity: a study utilizing Ecological Momentary Assessment. | Low RoB | Low RoB | High RoB | Low RoB | High RoB | High RoB | Low RoB | Low RoB | High RoB |
| Quinn 2014 | The impact of melancholia versus non-melancholia on resting-state, EEG alpha asymmetry: electrophysiological evidence for depression heterogeneity. | Low RoB | High RoB | Low RoB | Low RoB | High RoB | Low RoB | High RoB | Low RoB | Low RoB |
| Roh 2016 | EEG beta and low gamma power correlates with inattention in patients with major depressive disorder. | Low RoB | High RoB | N/a | N/a | Low RoB | Low RoB | High RoB | Low RoB | High RoB |
| Roh 2020 | Frontal Alpha Asymmetry Moderated by Suicidal Ideation in Patients with Major Depressive Disorder: A Comparison with Healthy Individuals. | Low RoB | Low RoB | Low RoB | Low RoB | Low RoB | Low RoB | Low RoB | Low RoB | High RoB |
| Saletu 2010 | EEG topography and tomography (LORETA) in diagnosis and pharmacotherapy of depression. | Low RoB | High RoB | Low RoB | High RoB | High RoB | Low RoB | High RoB | Low RoB | High RoB |
| Scanlon 2017 | Neurophysiologic Correlates of Headache Pain in Subjects With Major Depressive Disorder. | Low RoB | High RoB | N/a | N/a | Low RoB | High RoB | Low RoB | Low RoB | Low RoB |
| Segrave 2011 | Individualized alpha activity and frontal asymmetry in major depression. | Low RoB | Low RoB | Low RoB | Low RoB | High RoB | Low RoB | High RoB | Low RoB | Low RoB |

| | | | | | | | | | | |
|---|---|---|---|---|---|---|---|---|---|---|
| Soukhtanlou 2019 | Electrophysiological processing of happiness during conscious and sub-conscious awareness in depression | Low RoB | High RoB | Low RoB | High RoB | High RoB | High RoB | High RoB | Low RoB | Low RoB |
| Strelets 2007 | Comparative study of the gamma rhythm in normal conditions, during examination stress, and in patients with first depressive episode. | Low RoB | High RoB | High RoB | High RoB | High RoB | High RoB | High RoB | High RoB | High RoB |
| Tas 2015 | EEG power, cordance and coherence differences between unipolar and bipolar depression. | Low RoB | High RoB | N/a | N/a | High RoB | High RoB | High RoB | Low RoB | High RoB |
| Wu 2022 | Resting-state electroencephalography of neural oscillation and functional connectivity patterns in late-life depression. | Low RoB | High RoB | Low RoB | Low RoB | High RoB | Low RoB | High RoB | Low RoB | High RoB |
| Xia 2023 | Effects of Early Adverse Life Events on Depression and Cognitive Performance from the Perspective of the Heart-Brain Axis. | Low RoB | Low RoB | Low RoB | Low RoB | High RoB | High RoB | High RoB | Low RoB | High RoB |
| Zeng 2024 | Altered resting-state brain oscillation and the associated cognitive impairments in late-life depression with different depressive severity: An EEG power spectrum and functional connectivity study | Low RoB | High RoB | Low RoB | Low RoB | High RoB | High RoB | High RoB | Low RoB | Low RoB |
| Zhou 2023 | Alpha peak activity in resting-state EEG is associated with depressive score. | Low RoB | High RoB | N/a | N/a | High RoB | High RoB | High RoB | Low RoB | High RoB |
| Zoon 2013 | EEG alpha power as an intermediate measure between brain-derived neurotrophic factor Val66Met and depression severity in patients with major depressive disorder. | Low RoB | Low RoB | N/a | N/a | High RoB | High RoB | High RoB | Low RoB | Low RoB |

### Supplementary Figure S2:

Funnel plots for cross-sectional group comparisons:

**Delta power** (Egger's test p=0.3502)

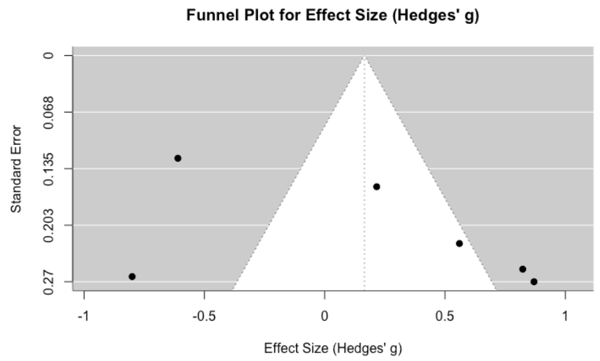


**Theta power** (Egger's test p=0.4085)

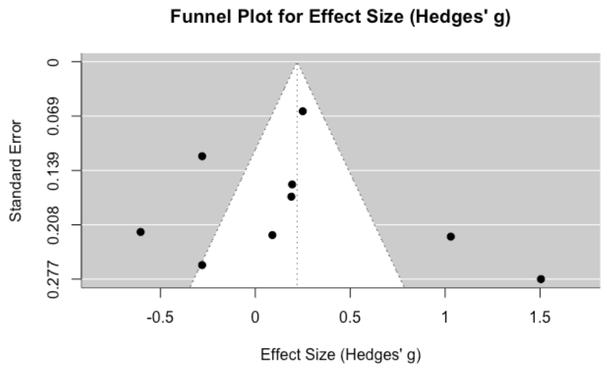

**Alpha power** (Egger's test p=0.0921)

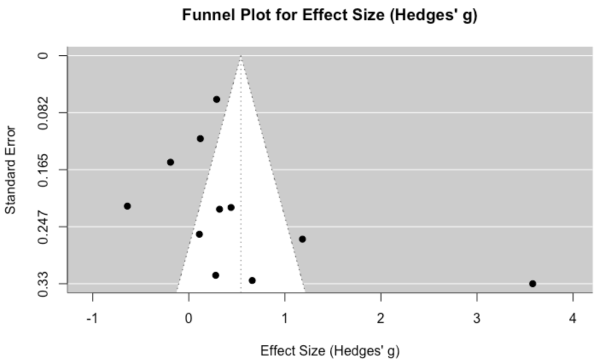


**Alpha Asymmetry** (Egger's test p=0.8254)

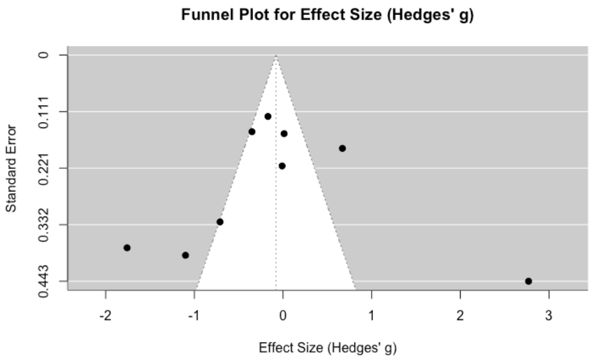


**Beta power** (Egger's test p=0.5407)

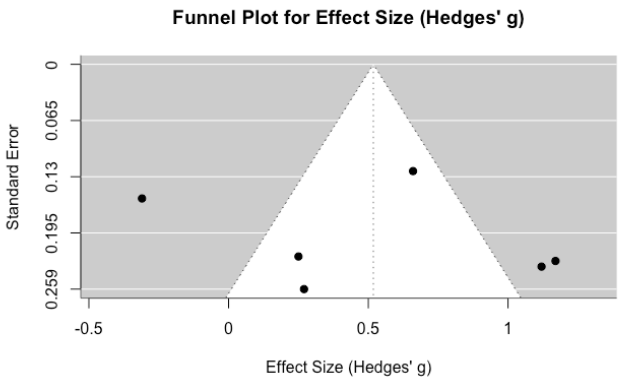

**Supplementary Figure S3:**

Funnel plots correlation analyses:

**Delta power** (Egger's test p=0.0573)

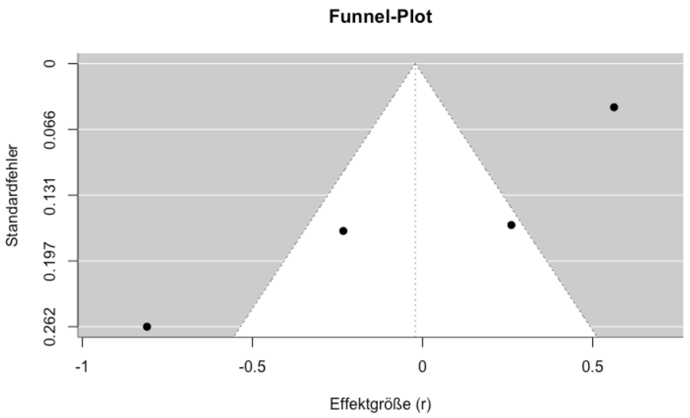


**Theta power** (Egger's test p=0.6960)

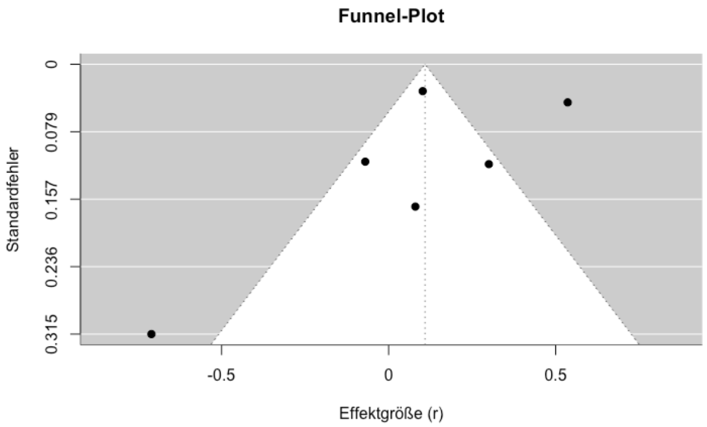


**Alpha power** (Egger's test p=0.2014)

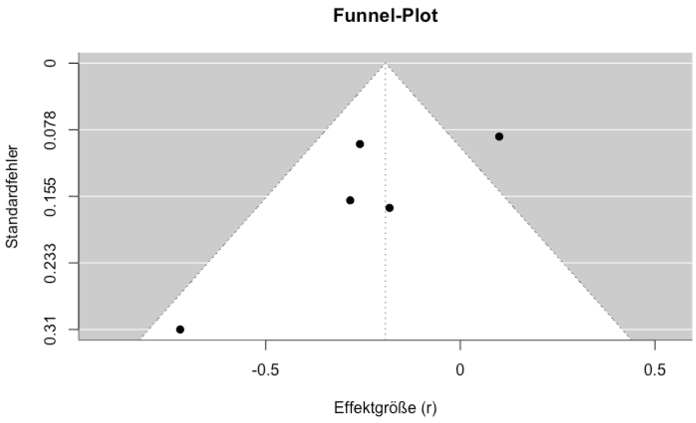

**Alpha Asymmetry** (Egger's test p=0.2548)

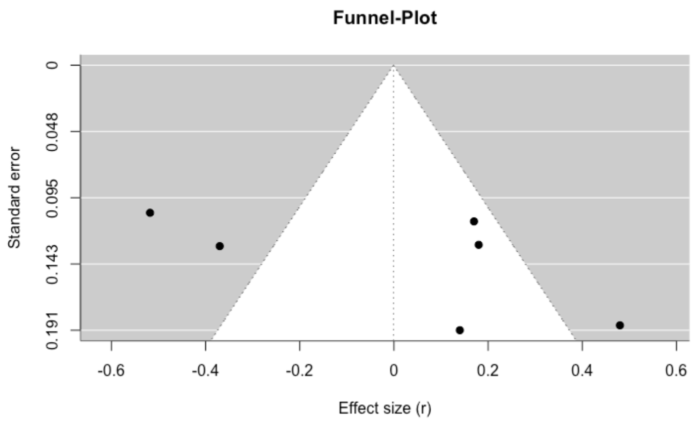


**Beta power** (Egger's test p=0.0673)

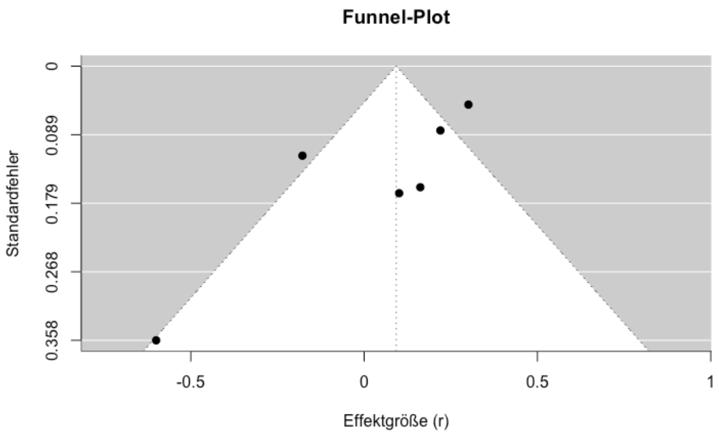
